# APOE-*ε4* Modulates Facial Neuromuscular Activity in Nondemented Adults: Toward Sensitive Speech-Based Diagnostics for Alzheimer’s Disease

**DOI:** 10.1101/2025.04.29.25326665

**Authors:** Marziye Eshghi, Panying Rong, Mehrdad Dadgostar, Hyunmin Shin, Brian D. Richburg, Nelson V. Barnett, David H. Salat, Steven E. Arnold, Jordan R. Green

## Abstract

The APOE-ε4 allele is a genetic risk factor for late-onset Alzheimer’s disease (AD). Beyond cognitive decline, APOE-ε4 affects motor function, reducing muscle strength and coordination, potentially through mitochondrial dysfunction and oxidative stress. This study examined the influence of the APOE-*ε4* allele on neuromuscular function in oral muscles involved in speech production, using surface electromyography (EMG); and assessed the predictive power of EMG measures in differentiating APOE-*ε4* carriers from noncarriers. Forty-two cognitively intact adults (16 APOE-*ε4* carriers, 26 noncarriers) completed speech tasks while EMG was recorded from seven craniofacial muscles. Seventy EMG features including amplitude, frequency, complexity, regularity, and functional connectivity were extracted. Statistical analyses assessed genotype effects, sex differences, and correlations with blood metabolic biomarkers. APOE-*ε4* carriers exhibited increased motor unit recruitment and synchronization, suggesting accelerated muscle fatigue. EMG-based measures outperformed cognitive tests in distinguishing carriers (AUC = 0.90) and correlated with metabolic biomarkers. Sex differences emerged, with female carriers showing reduced and male carriers showing increased functional connectivity. These findings highlight speech-based neuromuscular changes as potential early biomarkers of Alzheimer’s risk before cognition is affected.

## 1. Introduction

The Apolipoprotein E4 (APOE*-ε4*) allele is a well-established genetic risk factor for late-onset Alzheimer’s disease (AD), influencing disease progression and pathophysiology.^1,2^ This allele influences several biological markers of AD, including amyloid beta (Aβ) accumulation, cerebral glucose hypometabolism, cortical atrophy, and vascular abnormalities.^3–15^ Furthermore, APOE*-ε4* exacerbates tau pathology, neuroinflammation, and gliosis in both human AD brains and mouse models.^16–21^ Carriers of APOE-*ε4* face a significantly higher risk of developing mild cognitive impairment (MCI) and AD compared to non-carriers, with the presence of this allele potentially accelerating cognitive decline and leading to earlier onset of symptoms.^13,22–28^ Additionally, emerging research indicates that the APOE*-ε4* allele may significantly impact motor performance and neuromuscular function, leading to measurable changes in muscle strength, motor coordination, and overall physical performance.^29–36^ These effects suggest a broader influence of APOE*-ε4* beyond cognitive decline and AD risk, highlighting its potential role in neuromuscular degeneration and motor system vulnerabilities.

Research has shown that individuals carrying at least one copy of the *ε4* allele experience a faster rate of motor decline in older age, with a two-fold increase compared to non-carriers.^29^ This accelerated decline is largely attributed to changes in muscle strength, suggesting that APOE-*ε4* may influence neuromuscular function through its effects on muscular performance.^29^ APOE-*ε4* effects on motor function can be linked to APOE-*ε4*-mediated mechanisms such as mitochondrial dysfunction, oxidative stress, and disrupted synaptic signaling, all of which can impair motor performance and contribute to accelerated motor decline.^37^

While gross motor tasks primarily involve larger muscle groups and may overlook subtle deficits, speech production, which relies on highly coordinated, continuous activation of fine motor units, presents a unique opportunity to detect early motor impairments in APOE-ε4 carriers—potentially preceding overt cognitive or physical symptoms. Converging evidence shows that measurable subclinical changes in speech-related muscle activities can be detected early in neurodegeneration.^38–40^

This study aims to investigate the influence of the APOE-*ε4* allele on neuromuscular function in oral muscles involved in speech production, using surface electromyography (EMG) to capture muscle activity. The primary objective is to evaluate the predictive power of EMG measures extracted from craniofacial muscles during speech, comparing their accuracy in classifying APOE-*ε4* carriers and noncarriers with commonly used measures obtained from cognitive testing.

To understand the pathological mechanisms through which APOE-*ε4* impacts neuromuscular function, this study will further examine the associations between EMG measures and blood-based lipid and metabolic markers. These biomarkers will assess mitochondrial function, oxidative stress, and metabolic activity, shedding light on how these factors may influence orofacial neuromuscular functions in APOE-*ε4* carriers. Given the well-documented disruptive effects of APOE-*ε4* on mitochondrial pathways—causing oxidative stress, impaired ATP synthesis, and motor decline^29,37,41–43^—understanding the relationship between APOE-*ε4* pathology and neuromuscular dysfunction may provide novel insights into the early identification and monitoring of risk in APOE-*ε4* carriers.

We hypothesize that carrying the APOE-*ε4* allele contributes to motor unit remodeling or loss, resulting in reduced neuromuscular efficiency. These alterations could manifest as atypical motor unit recruitment and firing patterns during speech tasks, potentially reflected by increased complexity, greater variability, or the adoption of compensatory strategies. Such neuromuscular changes may be characterized by modified EMG signal amplitudes and frequency shifts, indicating altered conduction velocities and disrupted motor unit synchronization. Furthermore, the APOE-*ε4* allele may influence central motor pathways, affecting the functional connectivity between neural structures—such as brain-to-muscle interactions or coordination between functionally related muscle groups. These effects can be assessed using coherence measures obtained from EMG-EEG or EMG-EMG signal analyses. A reduction in coherence may suggest impaired neural signal integration during motor tasks. The neuromuscular alterations associated with APOE-*ε4* may stem from chronic metabolic impairments, contributing to motor unit remodeling, diminished neuromuscular efficiency, and accelerated muscle fatigue.

By investigating EMG patterns in speech-related muscles, this study aims to elucidate the impact of APOE-*ε4* on neuromuscular function, potentially identifying biomarkers useful for early diagnosis, disease monitoring, and the development of personalized therapeutic interventions.

## 2. Methods

The study was approved by the Mass General Brigham (MGB) Institutional Review Board (IRB Protocol #2021P001460). All procedures were conducted in accordance with MGB IRB guidelines and regulations. Written informed consent was obtained from all participants prior to their participation.

### 2.1 Participant population

Forty-two adults participated in this study. Participants were divided into two groups based on their APOE genotype: APOE*-ε4* positive (ε4+) (n=16) and APOE*-ε4* negative (ε4-) (n=26). The ε4+ group included individuals with ε3/ε4 heterozygous and ε4/ε4 homozygous genotypes, while the ε4-group consisted of individuals with the ε3/ε3 homozygous genotype. The groups were matched for age, and education (>12 years). *APOE* genotyping was conducted using blood samples, and all participants were blind to their respective genotypes. Of the 42 participants, 28 (7/5 female/male in the ε4+ group; 4/12 female/male in the ε4-group) had available blood-based lipid, metabolic, hematological, and vitamin/mineral markers, from which a subset was selected based on their relevance to mitochondrial function, metabolism, and oxidative stress. These data, along with demographic and clinical information for both groups are summarized in Table 1.

**Table 1.**
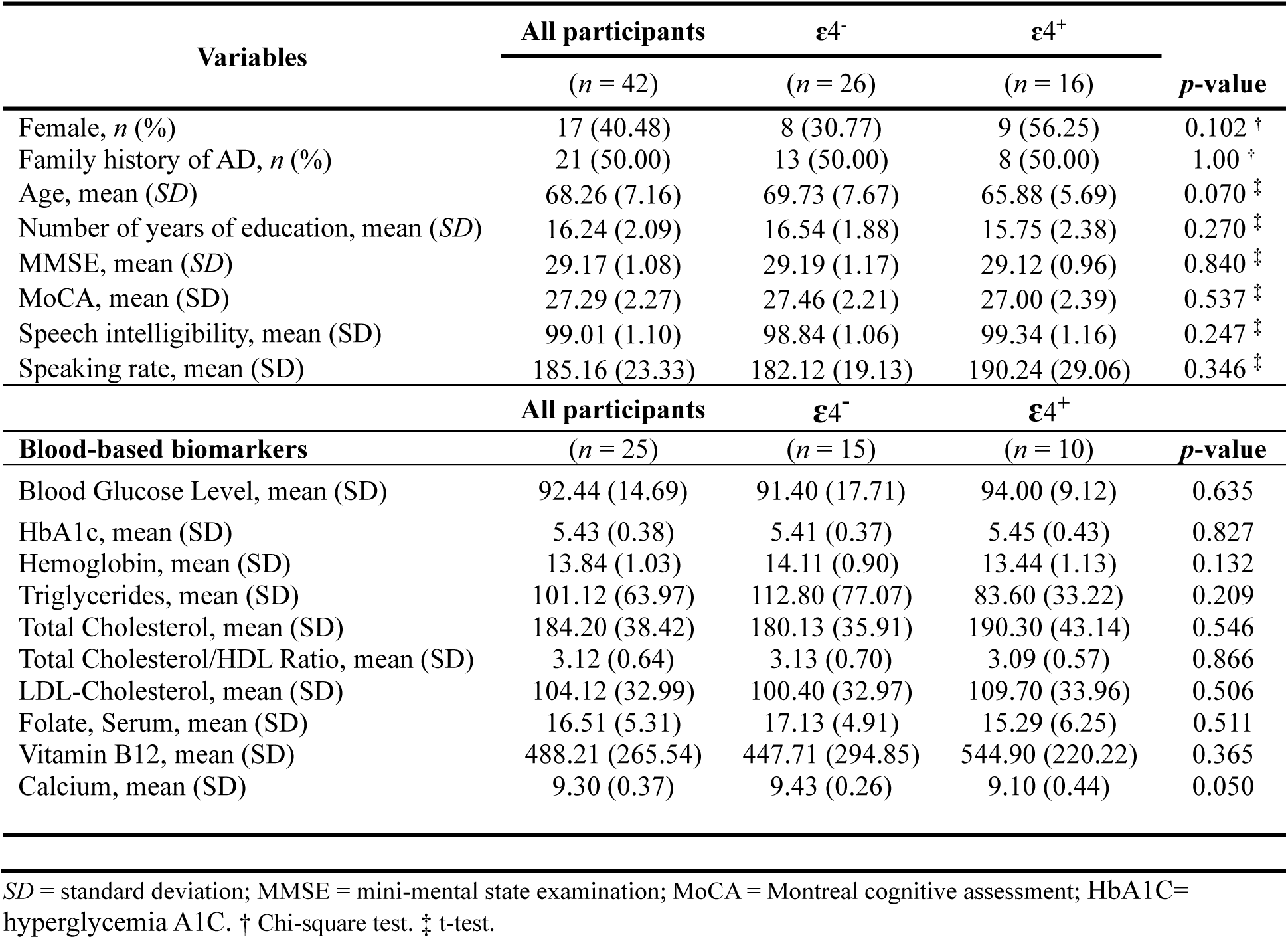
Demographic and clinical characteristics stratified by genotype carrier status.

All participants met the following inclusion criteria: (1) capacity to provide informed consent and comply with task-specific study instructions; (2) native speakers of American English; (3) no previous record of speech, language, hearing, psychiatric, or neurological disorders; (4) no clinical diagnosis of MCI; (5) not currently using psychoactive drugs that could potentially impact study outcomes; (6) passed an age-adjusted pure-tone hearing screening, ensuring normal hearing acuity in at least one ear; and (7) dental and occlusion status within functional limits, as determined by an oral examination performed by ME. Additionally, all participants demonstrated normal functional speech as assessed through the Sentence Intelligibility Test (SIT), a standardized tool that measures speech intelligibility and speaking rate. During the assessment, participants read 11 sentences randomly generated by the SIT software, with sentence length progressively increasing from 5 to 15 words. Their speech was digitally recorded and subsequently transcribed orthographically by two native American English-speaking listeners with established high interrater reliability (ICC= 0.99, p<0.001). The SIT software calculated speech intelligibility as the percentage of correctly transcribed words and speaking rate as the number of words read per minute (WPM). Furthermore, participants exhibited no sound distortions or abnormalities in voice, resonance, prosody, or rate qualities, as independently judged by two expert raters (ME and PR).

### 2.2 Experimental tasks

Participants were presented with nine sentences and instructed to read each sentence a fixed number of times. They produced five repetitions of the sentence “Buy Bobby a puppy” and two repetitions of each of the remaining eight sentences. These sentences were: “Billy blows big blue bubbles”; “Busy buzzing bumble bees”; “Bake big batches of bitter brown bread”; “Bankers pull plenty of bills into banks”; “Betty picked plenty of apple bits from the baby’s bib”; “Blue balloons pop when placed above sharp spikes”; “Ben’s brother, Patrick, beat up Ben’s pal, Bill”; and “Paul baked plenty of baguettes but put butter on the bottom.”

The sentences were selected because they include speech sounds that engage seven targeted speech muscles, including two lip muscles (inferior and superior orbicularis oris of the lip and five jaw muscles (left and right masseter, left and right temporalis, and anterior belly of the digastric (ABD)). The sentences are particularly rich in bilabial consonants (/b/, /p/, /m/), which require substantial activation of the orbicularis oris muscles. For instance, “Buy Bobby a Puppy” and “Billy blows big blue bubbles” prominently feature these sounds, making them ideal for observing the coordinated action of facial muscles. Similarly, the plosives (/b/, /p/) and fricatives (/f/, /v/) in phrases “Bake big batches” and “Blue balloons pop” engage the orbicularis oris, ABD and other facial muscles in rapid and precise movements. Alveolar sounds (/t/, /d/, /n/) in words like “Betty,” “plenty,” and “bottom” involve significant activation of the temporalis and masseter muscles for jaw positioning and stabilization. These sounds also recruit the ABD muscles for jaw opening, particularly in transitions between consonants and vowels. Phrases like “plenty” and “spikes” include consonant clusters that demand complex coordination across articulatory muscles, offering further insights into dynamic muscle control. The phonetic structure of sentences such as “Busy buzzing bumble bees” promotes repeated activation of the targeted craniofacial muscle groups. Additionally, the sentences feature a diverse range of vowel sounds, requiring varied configurations of the oral cavity and corresponding activation patterns of the speech muscles, including the ABD for jaw movement during low and mid vowels.^44^ While phonetically rich, these sentences also closely resemble natural speech, eliciting more realistic muscle activation patterns than those produced during isolated phoneme tasks. In summary, the combination of diverse consonant types and vowel contexts generates a comprehensive range of muscle activation patterns, making these stimuli particularly effective for EMG analysis of speech production.

### 2.3 Neuropsychological Assessment

Participants underwent a comprehensive array of standard neuropsychological tests routinely used to characterize AD-related conditions which assess attention, processing speed, learning, memory, language, and executive function. Cognitive tests used in this study included Montreal Cognitive Assessment (MoCA), Mini-Mental Status Examination (MMSE), Trail Making Tests A and B, the Hopkins Verbal Learning Test (HVLT), and the Controlled Oral Word Association Test (COWAT).

The *MoCA* is a widely used cognitive screening tool designed to evaluate a broad range of cognitive abilities and is particularly sensitive to detecting MCI and early-stage dementia.^45–50^ It provides a comprehensive assessment across multiple cognitive domains, including attention and concentration, executive functioning, memory, language, visuospatial abilities, orientation, and abstract thinking. Attention and concentration are assessed through digit spans, vigilance, and serial subtraction, while executive functioning is evaluated using tasks such as trail-making exercises and abstraction questions. Memory is measured through short-term recall of a word list, reflecting encoding and retrieval processes. Language is assessed through naming, sentence repetition, and verbal fluency tasks, while visuospatial abilities are tested using clock drawing and cube copying exercises. Orientation is evaluated through questions about the date, time, and place, and abstract thinking is measured by identifying similarities between items. The MoCA provides a total score out of 30, with lower scores indicating potential cognitive impairment. A score below the cutoff (typically 26) suggests the need for further assessment or intervention.

The *MMSE* is a widely used cognitive screening tool designed to assess global cognitive function and detect potential cognitive impairment.^51–57^ This brief, standardized test evaluates various cognitive domains, providing an overall measure of cognitive abilities. The MMSE assesses orientation by asking individuals to identify the date, day of the week, and their current location, reflecting awareness of time and place. Registration is tested by asking participants to repeat a list of three unrelated words, while attention and calculation are evaluated through tasks such as serial sevens (subtracting 7 repeatedly from 100) or spelling a word backward, measuring concentration and working memory. Delayed recall of the three words presented earlier assesses memory. Language abilities are tested through tasks like naming common objects, repeating sentences, following instructions, and writing a sentence, evaluating expressive and receptive language skills. Visuospatial skills are assessed by asking participants to copy a simple geometric figure, measuring visual perception and motor coordination. The MMSE provides a total score out of 30 points, with lower scores indicating greater cognitive impairment. A score of 24 or below is often used as a cutoff to indicate cognitive deficits, though adjustments may be made based on age, education, and cultural factors.

The *TMT*, comprising two parts (A and B), is a widely used neuropsychological assessment designed to evaluate various cognitive abilities, including visual attention, task switching, processing speed, and executive functioning.^58–64^ In Part A, participants connect a sequence of randomly arranged numbers in ascending order by drawing a line as quickly and accurately as possible. This segment primarily measures visual scanning, numerical sequencing, and motor speed. Part B requires participants to alternate between different cognitive tasks or mental sets, assessing cognitive flexibility, divided attention, and the inhibition of impulsive responses, while also incorporating the skills evaluated in Part A.^65–69^ Longer completion times and increased errors on the TMT are indicative of cognitive impairments and can affect everyday activities, including driving. Although the TMT is often scored using a ratio of Part B to Part A or by adjusting Part B scores relative to Part A, this analysis focused on evaluating each part independently. This approach allows for a more comprehensive exploration of distinct cognitive demands, encompassing both higher-order and lower-order processes.

The *HVLT* is a widely used measure of verbal learning and memory, consisting of multiple subtests that assess various aspects of cognitive functioning.^70–73^ These include immediate recall, delayed recall, and retention efficiency, which provide insights into encoding, storage, and retrieval processes in memory. The HVLT Immediate Total Recall subtest evaluates a participant’s ability to immediately recall a list of words presented verbally over multiple learning trials. It reflects short-term memory capacity, attention, and the initial encoding of verbal information. Performance on this measure is influenced by the ability to organize and retain information during the learning process. The HVLT Delayed Recall component assesses the participant’s ability to recall the same word list after a delay, without re-presentation of the list. It provides information on the strength of memory retention over time and the ability to retrieve information from long-term memory. Lower performance can indicate challenges with memory storage or retrieval. Finally, the HVLT Percent Retention measures the proportion of words recalled during the delayed recall phase relative to the words initially learned (immediate recall). It serves as an indicator of memory decay and retention efficiency. Higher retention percentages suggest more effective memory consolidation, while lower percentages may highlight difficulties in retaining learned information over time. These HVLT subtests, when analyzed together, offer a comprehensive understanding of verbal memory performance, enabling the identification of patterns associated with encoding, retention, and retrieval deficits.

The *COWAT*, specifically the FAS variant, is a widely used measure of phonemic verbal fluency designed to assess language production and executive functioning.^74–86^ This test examines the participant’s ability to generate words under specific phonological constraints, providing insights into cognitive processes such as mental flexibility, retrieval strategies, and inhibitory control. In this task, participants are asked to produce as many words as possible within a given time frame (typically 60 seconds per letter) that begin with a specified letter—F, A, or S—excluding proper nouns (i.e., name of people of places), numbers, and derivatives of the same word (i.e., repetitions of the same word with different affixes). Performance on this test evaluates the ability to organize and retrieve words based on phonemic rules while suppressing irrelevant or automatic responses. The FAS test relies on multiple cognitive domains, including language skills (retrieval of vocabulary and phonological processing), executive functioning (strategic planning, mental flexibility, and switching between subcategories within the letter constraint), and inhibitory control (suppressing inappropriate responses that do not meet the criteria). Reduced word generation or clustering, such as producing words only within a narrow subcategory, may indicate impairments in cognitive flexibility, processing speed, or access to lexical stores. Performance on the FAS test is commonly used in neuropsychological evaluations to identify deficits associated with frontal lobe dysfunction, neurological conditions, or disorders that impact executive functioning and language production.

All tests were scored using standardized norms adjusted for demographic variables including age, sex, education level, and ethnicity when appropriate to calculate the z-scores for each test. Subsequently, the cognitive composite z-score for each participant was calculated by summing the six standardized performance scores (i.e. TMT A, TMT B, HVLT-Immediate Total Recall, HVLT Delayed Recall, HVLT Percent Retention, COWAT-FAS). To evaluate the clinical diagnosis of MCI, participants’ cognitive status was assessed using five diagnostic frameworks: Conservative Subtype, Custom Criteria, Custom Subtype, Petersen Criteria, and Petersen Subtype. Approximately 60% of subjects demonstrated normal cognitive status across all criteria and subtypes, while 40% exhibited abnormal cognition (e.g., MCI, single amnestic) on two of these. However, all participants were ultimately classified as cognitively intact using a conservative approach, as they met the criteria for normal cognition in at least three of the five diagnostic frameworks. This method prioritizes higher specificity in identifying truly cognitively normal individuals. All neuropsychological data were administered, scored, and interpreted by an experienced neuropsychologist. Administration of neuropsychological batteries and EMG recordings took place either on the same day (for 12 participants) or on different days (for 30 participants), depending on the participant’s availability and preference. For sessions conducted on separate days, the interval was on average three months to maintain temporal proximity between cognitive and EMG data.

### 2.4 EMG Recording

EMG data collection was conducted in the Speech Physiology and Neurobiology of Aging and Dementia (SPaN-AD) lab at MGH Institute of Health Professions. Participants were seated in a chair, with speech tasks presented in a quasi-random order on a screen approximately 6 feet away. Each participant was fitted with surface electrodes for myoelectric data acquisition and a head-mounted microphone for acoustic data collection as described below.

EMG data were collected bilaterally from multiple facial and jaw muscles using a wireless EMG system (BIOPAC). Self-adhesive bipolar Ag/AgCl electrodes (15 mm diameter, 20 mm inter-electrode distance) were placed on the upper lip (two electrodes), lower lip (two electrodes), anterior belly of digastric (ABD, two electrodes), masseter (four electrodes, two pairs on each side), and temporalis (four electrodes, two pairs on each side). A ground electrode was attached behind the ear over the mastoid process.

Prior to electrode placement, the skin was prepared using an alcohol swab to increase conductance. Electrodes were positioned using craniofacial anatomical landmarks for reproducibility: along the cantho-gonial line for masseter, vertically at the coronal suture for anterior temporalis, and submentally along the posterior-inferior direction for ABD. Electrode placement was verified through calibration tasks such as jaw oscillations and clenching to ensure proper capture of muscle activities.

The EMG signals were pre-amplified by 1,000, band-pass filtered at 10-500 Hz, digitized at 10,000 Hz using the MP150 data acquisition module, and recorded using AcqKnowledge software (BIOPAC Systems, Inc.). Acoustic data used to annotate the EMG recordings were acquired using a Williams Sound MIC094 head-mounted microphone placed approximately 5 cm from the left lip corner. The audio signal was processed through a Focusrite Scarlett 2i2 audio interface and digitized at 10,000 Hz, synchronously with the EMG data. Participants were instructed to read sentences at their habitual speaking rate and loudness. The simultaneous recording of myoelectric and acoustic data allowed for temporal alignment during subsequent analysis.

### 2.5 EMG data processing and analysis

All EMG data processing and analytic procedures were implemented in MATLAB^87^ using custom-developed scripts. Following the best-practice guidelines for EMG signal processing,^88,89^ all seven channels were notch-filtered at 60 Hz and high-pass filtered at 20 Hz to remove power line noise and low-frequency motion artifacts, respectively. Lastly, the EMG signals for all repetitions of each stimulus were concatenated, resulting in 375 data samples (42 participants × 9 stimuli – 3 errors = 375) in total.

The processed EMG data samples were analyzed using an integrative automated analytic approach that adopted and combined two recently developed and validated EMG analytic algorithms, originally designed for a different neurodegenerative population.^38,90^ This method extracts 70 features across the seven EMG channels. An overview of these features is outlined in Table 2. This analytic approach combined linear and nonlinear analyses in time and frequency domains to measure the frequency, amplitude, complexity, and regularity of individual muscle activities, as well as the functional connectivity of the whole craniofacial muscle network.

**Table 2.**
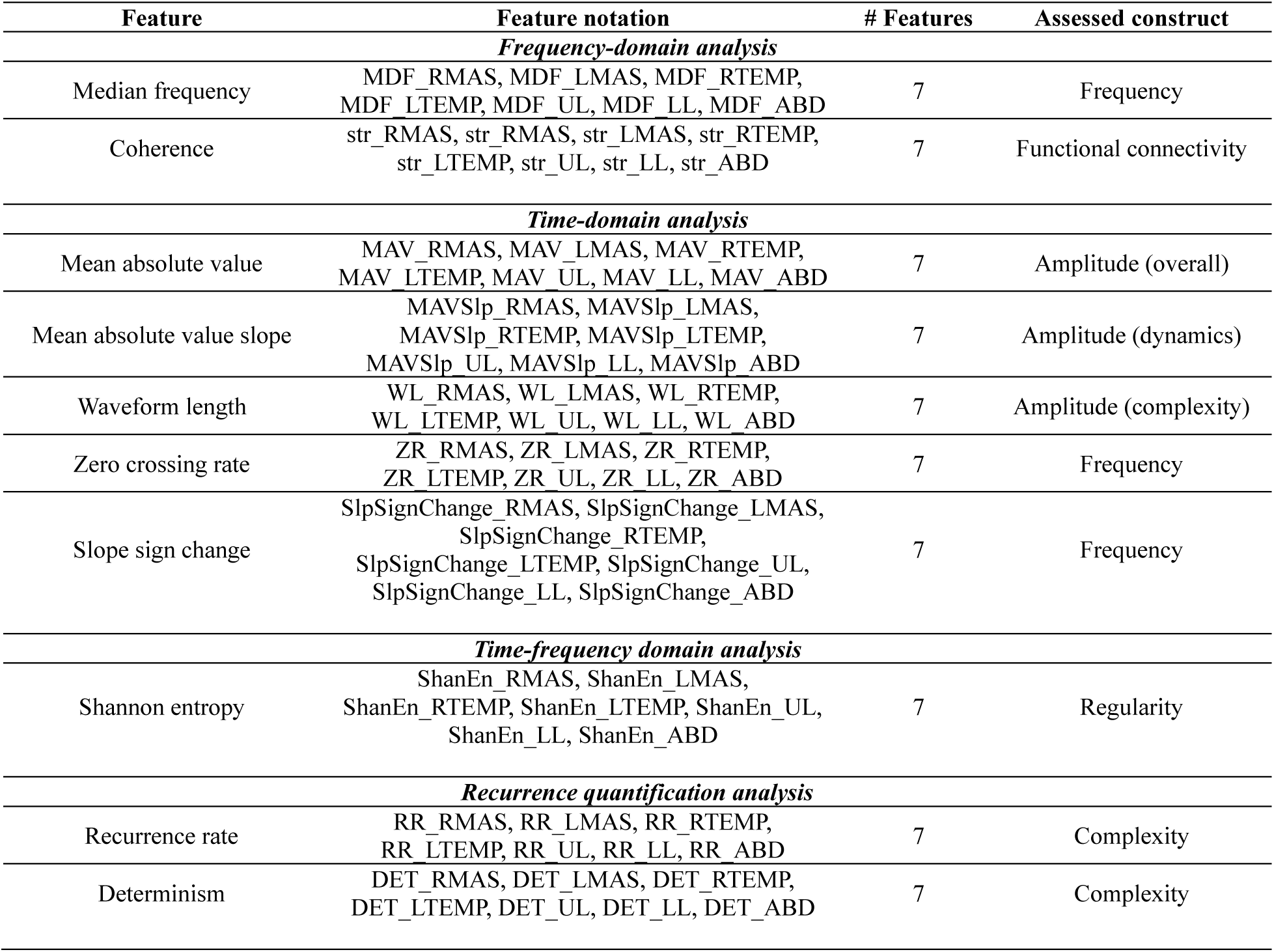
Overview of surface electromyography features.

| Feature | Feature notation | # Features | Assessed construct |
| --- | --- | --- | --- |
| *Frequency-domain analysis* |  |  |  |
| Median frequency | MDF_RMAS, MDF_LMAS, MDF_RTEMP,<br>MDF_LTEMP, MDF_UL, MDF_LL, MDF_ABD | 7 | Frequency |
| Coherence | str_RMAS, str_RMAS, str_LMAS, str_RTEMP,<br>str_LTEMP, str_UL, str_LL, str_ABD | 7 | Functional connectivity |
| *Time-domain analysis* |  |  |  |
| Mean absolute value | MAV_RMAS, MAV_LMAS, MAV_RTEMP,<br>MAV_LTEMP, MAV_UL, MAV_LL, MAV_ABD | 7 | Amplitude (overall) |
| Mean absolute value slope | MAVSlp_RMAS, MAVSlp_LMAS,<br>MAVSlp_RTEMP, MAVSlp_LTEMP,<br>MAVSlp_UL, MAVSlp_LL, MAVSlp_ABD | 7 | Amplitude (dynamics) |
| Waveform length | WL_RMAS, WL_LMAS, WL_RTEMP,<br>WL_LTEMP, WL_UL, WL_LL, WL_ABD | 7 | Amplitude (complexity) |
| Zero crossing rate | ZR_RMAS, ZR_LMAS, ZR_RTEMP,<br>ZR_LTEMP, ZR_UL, ZR_LL, ZR_ABD | 7 | Frequency |
| Slope sign change | SlpSignChange_RMAS, SlpSignChange_LMAS,<br>SlpSignChange_RTEMP,<br>SlpSignChange_LTEMP, SlpSignChange_UL,<br>SlpSignChange_LL, SlpSignChange_ABD | 7 | Frequency |
| *Time-frequency domain analysis* |  |  |  |
| Shannon entropy | ShanEn_RMAS, ShanEn_LMAS,<br>ShanEn_RTEMP, ShanEn_LTEMP, ShanEn_UL,<br>ShanEn_LL, ShanEn_ABD | 7 | Regularity |
| *Recurrence quantification analysis* |  |  |  |
| Recurrence rate | RR_RMAS, RR_LMAS, RR_RTEMP,<br>RR_LTEMP, RR_UL, RR_LL, RR_ABD | 7 | Complexity |
| Determinism | DET_RMAS, DET_LMAS, DET_RTEMP,<br>DET_LTEMP, DET_UL, DET_LL, DET_ABD | 7 | Complexity |

Technical details about these analyses are described below.

#### 2.5.1 Frequency-domain analysis

The frequency-domain analysis calculated the power spectral density of each EMG signal and the coherence spectra between different EMG signals. Power spectral density was estimated by Welch’s method, which was implemented by 1024-point Fast Fourier Transform (FFT) applied over a sliding 100-msec, 50% overlap Hamming window.^91^ The median frequency (MDF) of power spectral density was calculated as the frequency that divided the power spectrum into two parts with equal integrated spectral energy.

To calculate coherence spectra, all EMG signals were full wave rectified and reconstructed by concatenating stationary 1-sec epochs around the bursts for each channel. Magnitude-squared coherence spectrum was calculated for each pair of reconstructed signals, using 8192-point FFT applied over a sliding 8192-point Hamming window with 75% overlap, based on the Eq. 1:

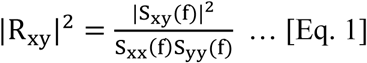

where |R_xy_|^2^ is magnitude-squared coherence spectrum, S_xy_(f) is the cross-spectrum between two muscles, and S_xx_(f) and S_yy_(f) are the auto-spectra for each muscle. In addition, a significance level corresponding to the upper 95% confidence limit under the hypothesis of independence between muscles was calculated as: *S* = 1 − 0.05^1/(*L^*−1)^, where *L*^ is the adjusted number of overlapped segments in coherence calculation.^92,93^ The significance level was used to exclude spurious functional connections between muscles.

In this study, we focused on the coherence in the beta band. Beta oscillations (15-35 Hz) originate from the motor cortical network, providing a descending drive for synchronous modulation of functionally related muscles.^94–96^ Thus, beta-band coherence can be interpreted as a measure of functional connectivity between muscles related to a common cortical drive. To measure beta-band coherence, the significance of the coherence spectrum within the beta band was first verified. Specifically, if no peak in the beta band of the coherence spectrum exceeded the significance level, the functional connection between the muscles was regarded as spurious, and beta-band coherence was set to zero; otherwise, the mean coherence within the beta band was calculated. After the verification of significance, all beta-band coherence metrics were transformed to Fisher z-scores for variance stabilization.^92^ Lastly, the mean strength of functional connectivity was calculated for each muscle as the average sum of Fisher z-score of beta-band coherence between this and all other muscles.

#### 2.5.2 Time-domain analysis

The time-domain analysis was applied to 100-msec segments centered at the bursts, and the features extracted were averaged across all segments for each channel. Such short segments represent the initial phase of muscle contraction, which encodes the most stable and useful neuromuscular information.^97^ This analysis extracted five features for each muscle, including:

1. Mean absolute value (MAV), defined as the mean absolute value of all data points within a segment:

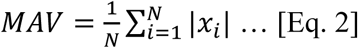

where N is the number of data points in the segment and *x*_*i*_ is i^th^ data point in the segment.
2. Mean absolute value slope (MAVSlp), defined as the difference of MAV between adjacent segments:

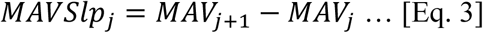

where *j* = 1,2, …, *J* − 1 and *J* is the total number of segments in the EMG signal.
3. Zero crossings (ZR), defined as the number of times the waveform crosses zero:

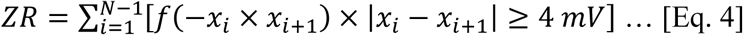

where 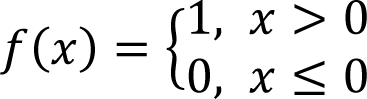. To eliminate noise fluctuations, a 4 *mV* threshold (i.e., 4 μ*V* peak-to-peak intrinsic system noise × 2000 gain/2) is implemented while calculating ZR.
4. Slope sign change (SlpSignChange), defined as the number of times the slope of the waveform changes sign:

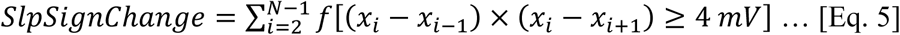 Like for ZR, a 4 *mV* threshold is employed to reduce noise-induced slope sign changes.
5. Waveform length (WL), defined as the cumulative length of the waveform over the segment:

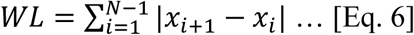

#### 2.5.3 Time-frequency domain analysis

First, each EMG signal was decomposed into eight scale-frequency representations, using 3-level wavelet packet decomposition with the 5^th^ order Symlets wavelet.^98^ Next, the Shannon entropy of each time-frequency representation was computed as:

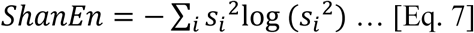

where *s*_*i*_ is the i^th^ coefficient of the scale-frequency representation, reflecting the approximation (i.e., low-pass) or detail (i.e., high-pass) coefficients at each decomposition level. Lastly, the sum of the Shannon entropy of all eight scale-frequency representations was calculated to index the irregularity of myoelectric activity for each muscle.

#### 2.5.4 Recurrence Quantification Analysis (RQA)

RQA quantified the structural complexity of EMG signals based on the dynamics of state change in the recurrence plot.^99,100^ A recurrence plot is composed of recurrence points, where a recurrence point is defined as a state *j* when its phase space trajectory *x⃗*_*j*_ returns to a previous state *i* so that the distance between *x⃗*_*j*_ and *x⃗*_*i*_ is below the threshold ε. Here the phase space trajectory *x⃗*_*j*_ is defined as: *x⃗*_*j*_ = [*u*_*j*_, *u*_*j*+τ_, … *u*_*j*+(*m*−1)τ_], where *m* is the embedding dimension parameter, and τ is the time delay parameter. Recurrent plots were generated for non-overlapping 1-sec segments of each EMG signal, with the parameters set to the following pre-determined values: *m* = 30, τ = 3, ε = 0.2. Such segments represented stationary intervals of EMG signals, allowing for reliable analysis of the underlying motor unit firing patterns. Two quantitative metrics—recurrence rate (RR) and determinism (DET)—were derived from each segment and then averaged across all segments for each muscle. RR measured the ratio of all recurrence points to all possible states in the recurrence plot. DET measured the ratio of recurrence points forming diagonal structures to all recurrence points, which can be interpreted as an index of the overall periodic content in the signal.

### 2.6 Statistical analysis

All statistical analyses were conducted in the R statistical computing program,^101^ with the significance level set to *p* < .05. Prior to the analyses, all EMG features were subject to a data cleaning and imputation procedure, by (1) identifying and excluding all outliers outside the range of [*lower quantile – 1.5* IQR, upper quantile + 1.5*IQR*] and (2) imputing missing data using the k-nearest neighbors (KNN) algorithm (*preProcess*).^102^ The imputed data were submitted to statistical analyses as described below.

#### 2.6.1 Effect of APOE-ε4 on neuromuscular function

To provide an initial inspection of the effect of APOE*-ε4* on neuromuscular function, the effect size of the difference between all EMG features for APOE*-ε4* carriers and noncarriers was calculated using Cohen’s d. Due to the differential impact of APOE*-ε4* on males and females,^103–107^ the effect sizes were calculated for the two sexes separately.

#### 2.6.2 Factor analysis

To reduce the dimension of the EMG feature set, all features were submitted to maximum likelihood factor analysis (*factanal*) with *oblimin* rotation to cluster features that shared common variance into a lower-dimensional set of latent factors, where the number of factors was determined by parallel analysis (*fa.parallel*).^108^ This procedure aimed to circumvent the curse of dimensionality, as commonly encountered in digital health applications,^109^ to avoid overfitting to a specific dataset in the subsequent classification analysis. The resulting latent factors represented unobserved neuromuscular traits of the craniofacial muscle network. Factor scores were estimated using the *tenBerge* method, providing EMG-based measures of participants’ neuromuscular performance in the latent factor space.

#### 2.6.3 Predictive power of EMG-based measures for classifying APOE-ε4 carriers from noncarriers

To evaluate the discriminatory power of each EMG-based measure for differentiating APOE*-ε4* carriers from noncarriers, each EMG-based measure was submitted to a linear mixed effects model, with genotype (i.e., binary variable denoting APOE*-ε4* carriers versus noncarriers), sex, age, and the interaction between genotype and sex as the fixed effects, and a participant-dependent intercept term as the random effect. The main effect of APOE*-ε4* and its interaction with sex were the primary effects of interest for evaluating the significance of carrier-noncarrier difference. Next, all EMG-based measures were aggregated by averaging across the nine stimuli for each participant. The Receiver Operating Characteristic (ROC) statistic was calculated for each aggregated measure to index its discriminatory power for differentiating carriers from noncarriers.

To compare the discriminatory power of the EMG-based measures with standard measures derived from the cognitive domain (i.e., scores on MoCA,MMSE, and the comprehensive z-score of all domain-specific neuropsychological tests), all cognitive measures underwent a similar analysis as above. Specifically, general linear models were used to evaluate the effects of genotype, sex, age, and the interaction between genotype and sex on each cognitive measure. Furthermore, the ROC statistic was calculated for all cognitive measures.

In addition to the discriminatory power of individual measures, we also evaluated the efficacy of the combination of all EMG-based measures for differentiating APOE*-ε4* carriers from noncarriers. To this end, all EMG-based measures, along with age and sex, were fed into penalized logistic regression with the elastic net penalty to classify between the samples for carriers and noncarriers. The penalty was imposed to reduce the coefficients of minimally contributing predictors toward zero, allowing the coefficients to inform the relative importance of the predictors. The performance of classification was estimated by the area under the curve (AUC), accuracy, sensitivity, and specificity, through 5-fold cross-validation repeated 10 times.

#### 2.6.4 Correlation of EMG-based measures with blood-based biomarkers

Pearson’s correlation coefficient was calculated between all EMG-based measures and blood-based biomarkers. The magnitude of the correlation coefficients was interpreted to evaluate the relation between the biolological and neuromuscular factors, in order to identify potential biological underpinnings of the APOE*-ε4*-induced neuromuscular changes.

## 3. Results

### 3.1 Effect of APOE-ε4 on EMG features

The effect size of the difference between the EMG features for APOE-*ε4* carriers and noncarriers is displayed by sex in Figure 1. Based on these results, the APOE-*ε4* allele showed both common and distinct effects on neuromuscular function for males and females.

**Figure 1.**
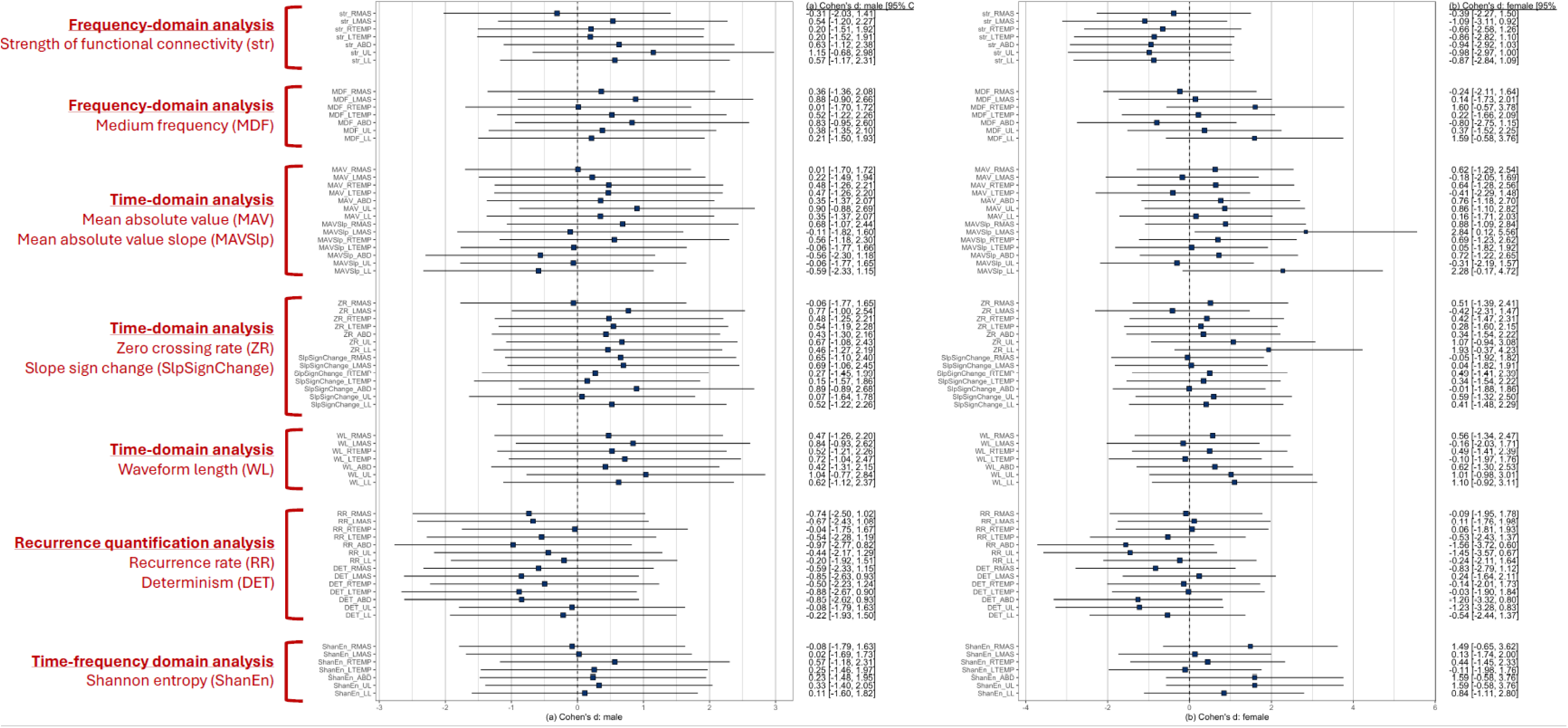
Cohen’s d effect size of the difference between the surface electromyograph features for (a) male APOE-ε4 carriers and noncarriers; (b) female APOE-ε4 carriers and noncarriers. RMAS = right masseter, LMAS = left masseter, RTEMP = right temporalis, LTEMP = left temporalis, ABD = anterior belly of digastric, UL = upper lip muscles, LL = lower lip muscles.

Focusing on the features that exhibited at least a medium effect size (|d| > 0.50), the common effects of APOE-*ε4* for both sexes included (1) increased amplitude (i.e., increased MAV and WL), especially for lip muscles, (2) increased frequency (i.e., increased MDF, ZR, SlpSignChange), (3) increased complexity (i.e., decreased RR and DET), and (4) decreased regularity (i.e., increased ShanEn). The distinct effects of APOE-*ε4* for males and females were primarily manifested in functional connectivity. Specifically, male APOE-*ε4* carriers exhibited a trend toward increased functional connectivity (i.e., increased str), whereas female APOE-*ε4* carriers showed an opposite trend toward reduced functional connectivity (i.e., decreased str), relative to their noncarrier counterparts.

### 3.2 Factor analysis

The 70 EMG features were clustered into 14 latent factors. The resulting factorization model accounted for 64.1% of the total variance in the original feature set and was determined to be sufficient based on χ^2^ test (χ^2^ = 4298.64, *p* < .001). The rotated factor loadings are shown in Figure 2.

**Figure 2.**
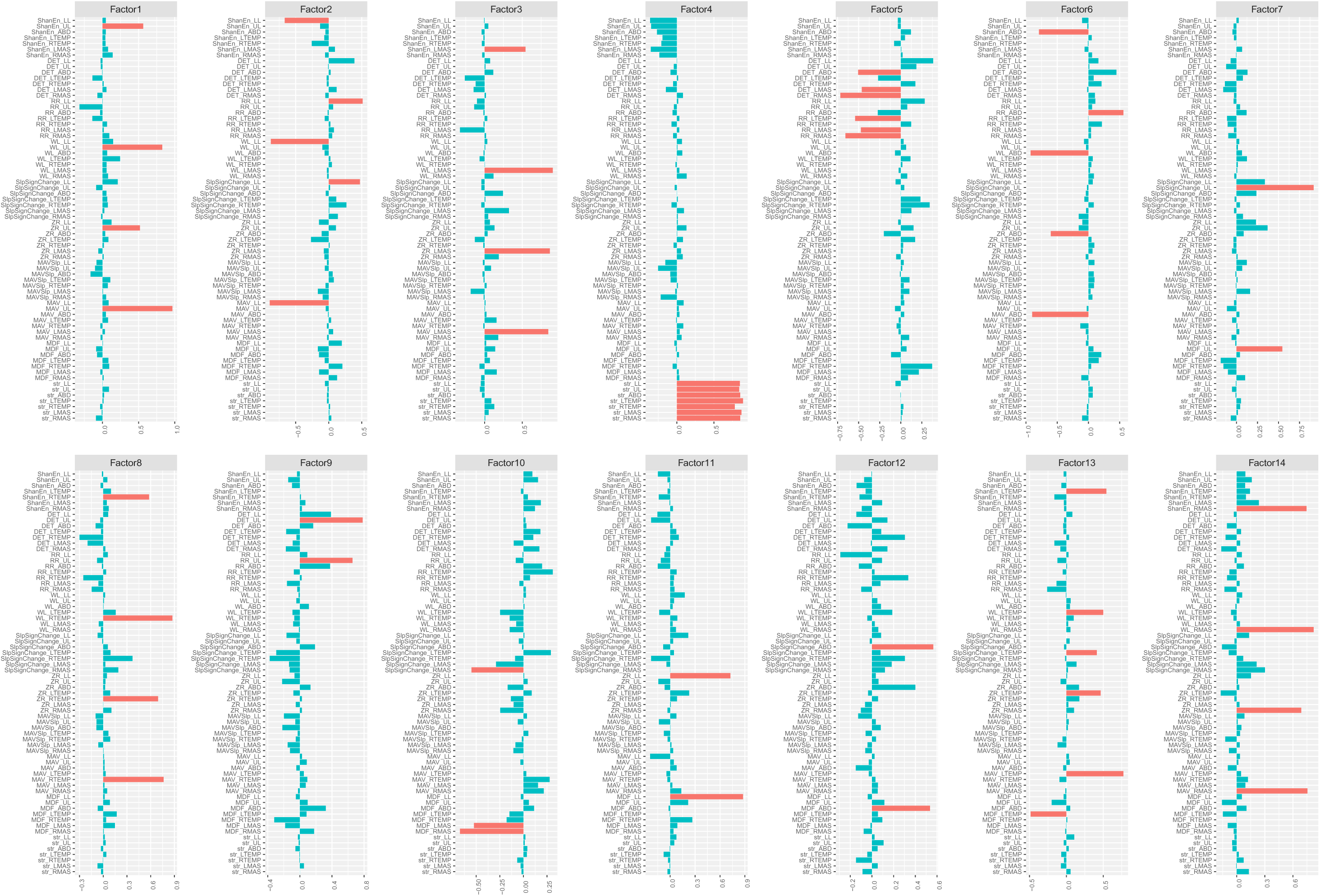
Rotated factor loadings. The height of the bars represents the loading of the surface electromyography features on each latent factor. By convention, features that load greater than 0.4 (i.e., absolute value > 0.4) are regarded as the primary component features of the latent factor, and the loadings of these features are shown in red, while the loadings of the other features are shown in blue.

Features that loaded greater than 0.4 (i.e., absolute factor loading > 0.4) were conventionally regarded as the primary component features of the factor. Accordingly, the primary component features of factors 1 through 14 are identified and listed in Table 3. Based on these feature clusters, all 14 latent factors were interpretable, reflecting either muscle-specific (i.e., factors 1, 2, 3, 6, 8, 13, 14) or construct-specific (i.e., factors 4, 5, 7, 9, 10, 11, 12) characteristics.

**Table 3.** Results of factor analysis.

| **Factor** | **Latent traits** | **Primary component features** |
| --- | --- | --- |
| X1 | Global characteristics of upper lip muscles | amplitude (MAV, WL), regularity (ShanEn), frequency (ZR) |
| X2 | Global characteristics of lower lip muscles | amplitude (MAV, WL), complexity (RR), regularity (ShanEn), frequency (SlpSignChange) |
| X3 | Global characteristics of left masster | amplitude (MAV, WL), regularity (ShanEn), frequency (ZR) |
| X4 | Global functional connectivity of all muscles | functional connectivity (str) |
| X5 | Complexity of jaw muscles | complexity (RR, DET) |
| X6 | Global characteristics of ABD | amplitude (MAV, WL), complexity (RR), regularity (ShanEn), frequency (ZR) |
| X7 | Frequency of upper lip muscles | frequency (MDF, SlpSignChange) |
| X8 | Global characteristics of right temporalis | amplitude (MAV, WL), regularity (ShanEn), frequency (ZR) |
| X9 | Complexity of upper lip muscles | complexity (RR, DET) |
| X10 | Frequency of masseters | frequency (MDF, SlpSignChange) |
| X11 | Frequency of lower lip muscles | frequency (MDF, ZR) |
| X12 | Frequency of ABD | frequency (MDF, SlpSignChange) |
| X13 | Global characteristics of left temporalis | amplitude (MAV, WL), regularity (ShanEn), frequency (MDF, ZR, SlpSignChange) |
| X14 | Global characteristics of right masster | amplitude (MAV, WL), regularity (ShanEn), frequency (ZR) |

### 3.3 Discriminatory power of EMG-based measures for classifying APOE-ε4 carriers from noncarriers

Of all 14 EMG-based measures, sex showed a significant effect on one muscle-specific measure (i.e., X6, reflecting ABD characteristics), *F*(1,37.01) = 5.08, *p* = .027. Genotype, age, and the interaction between genotype and sex did not have a significant effect on any of the measures. Figure 3 shows the box plots for all EMG-based measures by genotype and sex. The ROC statistics, including AUC and the sensitivity and specificity at the optimal cutoff that maximizes the sum of sensitivity and specificity, are provided in Table 4.

**Figure 3.**
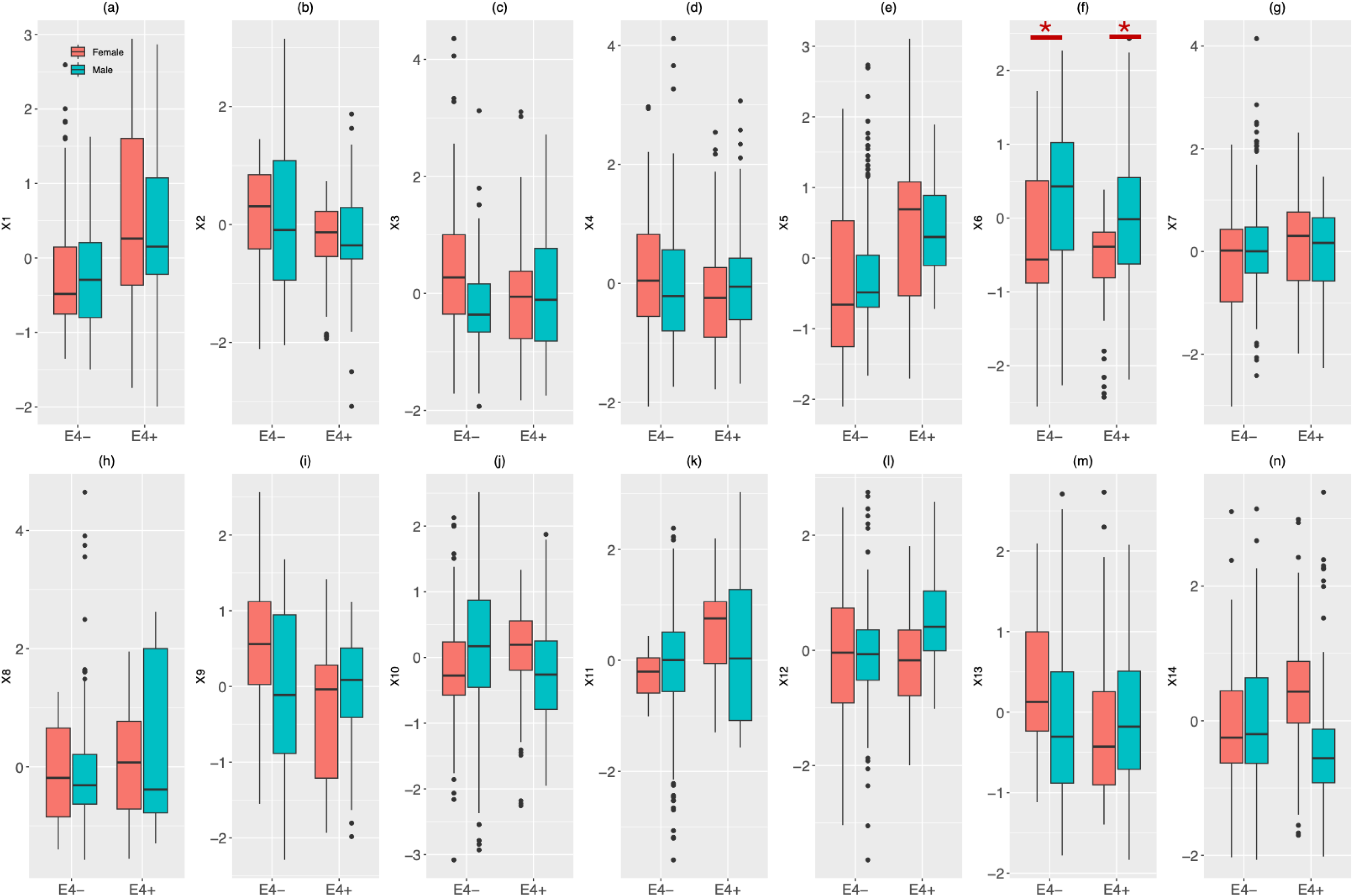
Box plots for EMG-based measures (X1 – X14) by genotype (wE4: APOE-ε4 carriers; woE4: APOE-ε4 noncarriers) and biological sex.

**Table 4.** Area under the curve (AUC), sensitivity, and specificity of all EMG-based measures (X1 – X14) and cognitive measures (CompCog: composite z-score of all domain-specific neuropsychological tests; MoCA: z-score on the Montreal Cognitive Assessment; MMSE: z-score on the Mini-Mental State Exam) for differentiating APOE-*ε4* carriers from noncarriers. The last column shows the coefficients of penalized logistic regression, reflecting the relative importance of the EMG-based measures to the classification model between carriers and noncarriers.

| Measure | AUC | Sensitivity | Specificity | Coefficient |
| --- | --- | --- | --- | --- |
| X1 | 0.70 | 0.81 | 0.60 | 0.73 |
| X2 | 0.60 | 0.48 | 0.93 | -0.65 |
| X3 | 0.51 | 0.67 | 0.47 | 0.28 |
| X4 | 0.54 | 0.67 | 0.53 | -0.058 |
| X5 | 0.68 | 0.74 | 0.73 | 0.45 |
| X6 | 0.59 | 0.52 | 0.87 | -0.67 |
| X7 | 0.57 | 0.89 | 0.40 | 0.35 |
| X8 | 0.53 | 0.56 | 0.60 | 0.39 |
| X9 | 0.60 | 0.44 | 0.93 | -0.33 |
| X10 | 0.55 | 0.19 | 1.00 | -0.87 |
| X11 | 0.67 | 0.89 | 0.53 | 1.39 |
| X12 | 0.57 | 0.67 | 0.53 | 0.62 |
| X13 | 0.58 | 0.70 | 0.53 | -0.68 |
| X14 | 0.54 | 0.81 | 0.40 | 0.091 |
| CompCog | 0.48 | 0.63 | 0.53 | - |
| MoCA | 0.49 | 0.26 | 0.80 | - |
| MMSE | 0.61 | 0.30 | 0.93 | - |

Of the three standard cognitive measures, no significant effect was found for genotype, sex, or the interaction between genotype and sex, whereas age showed a significant effect on the z-score of MMSE, *F*(1,37) = 14.32, *p* < .001. The ROC statistics for all cognitive measures are displayed alongside the EMG-based measures in Table 4. Based on the comparisons of the ROC statistics, the best-performing EMG-based and cognitive measures were identified, which were X1 and the z-score of MMSE, respectively, and their ROC curves are shown in Figure 4.

**Figure 4.**
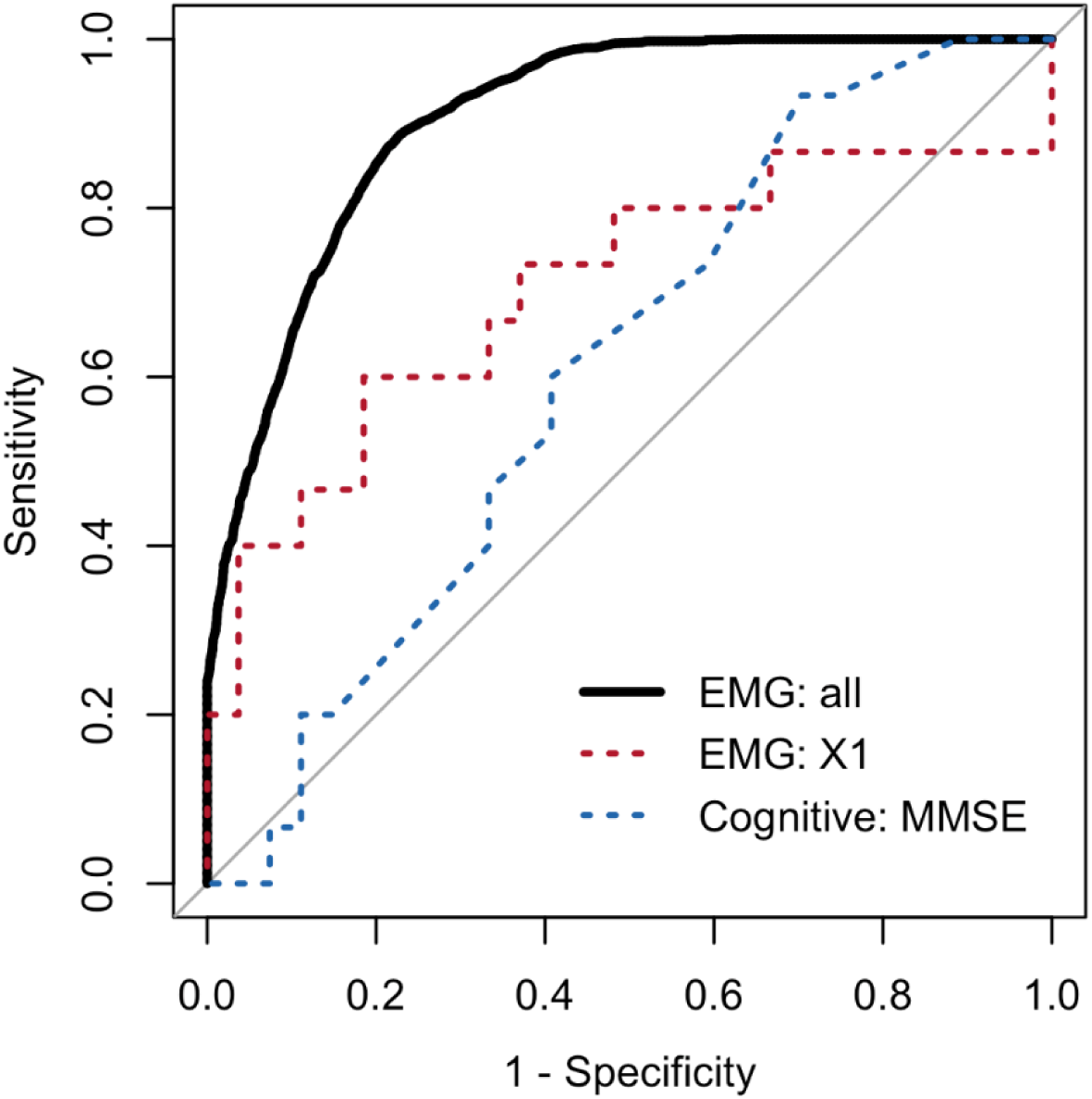
Receiver Operating Characteristic (ROC) curves for (1) the best-performing EMG-based measure (i.e., X1) and cognitive measure (i.e., z-score on the Mini-Mental State Exam [MMSE]) for differentiating APOE-ε4 carriers from noncarriers, out of all 14 EMG-based measures and three cognitive measures, and (2) the combination of all 14 EMG-based measures (i.e., X1-X14), based on multivariate classification using penalized logistic regression.

The classification model based on all EMG-based measures showed an AUC of 0.91, with 0.82 accuracy, 0.88 sensitivity, and 0.70 specificity, for differentiating APOE-*ε4* carriers from noncarriers. The coefficients of the predictors, reflecting the relative importance of each EMG-based measure to the outcome, are shown in Table 4. In addition to these EMG-based measures, age also comprised an important contributor to the classification (coefficient = -1.45), whereas the contribution of sex was more trivial (coefficient = 0.50).

Taken together the ROC statistics for individual EMG measures and the relative importance of these measures in the multivariate classification model, X1, X2, and X11 appear to have overall the greatest discriminatory power for differentiating APOE-*ε4* carriers from noncarriers (AUC ≥ 0.60; |coefficient| ≥ 0.65). These measures included two muscle-specific metrics (X1, reflecting the upper lip muscle characteristics; X2, reflecting the lower lip muscle characteristics) and one construct-specific metric (X11, reflecting the frequency of lower lip muscles).

### 3.4 Correlation of EMG-based measures with blood-based biomarkers

For the EMG-based measures with the highest discriminatory power in classifying APOE-ε4 carriers versus noncarriers (i.e., X1, X2, X11), the corresponding biological correlates were identified as those biomarkers exhibiting an absolute correlation coefficient greater than 0.3, indicating a large effect size.^110^ Accordingly, the biological correlates of X1 included the total and LDL cholesterol. An increase of X1 (reflecting increased amplitude, irregularity, and frequency of upper lip muscles, as observed in APOE-*ε4* carriers from Figures 1 and 3) was correlated with decreases in total and LDL cholesterol. The biological correlates of X2 included glucose, hemoglobin, total and LDL cholesterol, as well as vitamin B12. A decrease of X2 (reflecting increased amplitude, irregularity, and frequency of upper lip muscles, as observed in APOE-*ε4* carriers from Figures 1 and 3) was correlated with decreased glucose, hemoglobin, total and LDL cholesterol and increased vitamin B12. The biological correlates of X11 were triglycerides, vitamin B12, and serum folate. An increase of X11 (reflecting increased frequency of lower lip muscles, as observed in APOE-*ε4* carriers from Figures 1 and 3) was correlated with decreased triglycerides and increased vitamin B12 and serum folate.

## 4. Discussion

This study investigated the influence of a known genetic risk factor for AD – APOE*-ε4* – on the neuromuscular function of speech-related muscles. Features extracted from craniofacial muscles outperformed cognitive measures in classifying APOE*-ε4* carriers from noncarriers. We identified two potential neuromuscular risk factors for AD:increased motor unit recruitment and synchronization. These risk factors are likely associated with accelerated muscle fatigue, driven by APOE-*ε4*-mediated changes in the central nervous system and neuromuscular function, potentially linked to mitochondrial dysfunction, oxidative stress, and impaired cellular metabolism.

### 4.1 Speech-based neuromuscular risk factors for AD

Although APOE-*ε4* is a known risk factor for cognitive decline and AD, its effects on motor function remain underexplored. This study reveals altered neuromuscular function in APOE-*ε4* carriers during speech, marked by increased amplitude, frequency, complexity, and irregularity of myoelectric signals across facial and jaw muscles (Figure 1). These patterns suggest a shift in motor unit recruitment strategies, characterized by increased activation of larger, fast-conducting motor units and greater synchronization of smaller motor units. Both are well-documented markers of neuromuscular fatigue,^91,111^ potentially reflecting a compensatory response to early metabolic inefficiencies or motor unit remodeling associated with APOE-*ε4*. This adaptation may help sustain force production but could also contribute to premature fatigue and altered motor control dynamics. These adaptations may also lead to higher frequency of muscle activity^112^ and increased heterogeneity in the physiological properties of recruited motor unit pools, resulting in more complex and irregular firing patterns.

### 4.2 Biological correlates of neuromuscular changes

The link between APOE-*ε4*-associated neuromuscular changes and muscle fatigue is reinforced by biochemical correlates of EMG features. Key distinguishing factors (X1: global characteristics of upper lip muscles; X2: global characteristics of lower lip muscles; X11: frequency of lower lip muscles) correlate with total and LDL cholesterol, triglycerides, hemoglobin, glucose, vitamin B12, and serum folate. These biomarkers have critical implications for mitochondrial function and oxidative stress.

Cholesterol and triglycerides shape mitochondrial membranes and influence energy production and oxidative stress^113^— processes often dysregulated in APOE-*ε4* carriers due to altered lipid metabolism.^114,115^ The accumulation of these metabolites can lead to increased group III and IV afferent feedback, necessitating compensatory increases in motor unit recruitment and discharge rates to maintain task demands.^116–118^ Vitamin B12 and folate play essential roles in one-carbon metabolism and homocysteine regulation, affecting DNA synthesis, methylation reactions, and overall cellular energy balance.^119–121^ Deficiencies in these vitamins can lead to elevated homocysteine levels, a risk factor for cognitive decline, and potentially contribute to the accelerated muscle fatigue observed in APOE-*ε4* carriers.^113^ Finally, hemoglobin is essential for oxygen transport and mitochondrial ATP production.^121,122^ Impaired oxygen delivery can shift metabolism toward anaerobic glycolysis, leading to lactate buildup and muscle fatigue.^113^

### 4.3 Cellular mechanisms underlying neuromuscular dysfunction in APOE-ε4 carriers

At the molecular level, APOE-*ε4* disrupts mitochondrial pathways involved in electron transport, ATP synthesis, and thermogenesis,^123^ contributing to accelerated muscle fatigue and motor decline. Moreover, APOE-*ε4* carriers exhibit heightened oxidative stress, particularly in synaptosomal membranes, leading to neuronal damage, motor neuron degeneration, and astrocyte dysfunction.^123–129^ The interplay of oxidative stress, mitochondrial inefficiency, and disrupted synaptic signaling might have collectively influenced neuromuscular function in APOE-*ε4* carriers in the current study, aligning with previous research.^29,130,131^

Separately, the APOE-*ε4* allele significantly impacts calcium ion dynamics and excitation-contraction coupling, potentially disrupting motor unit recruitment and synchronization. Studies have linked APOE-*ε4* to mitochondrial calcium overload, demonstrated in Neuro-2A cells and rat hippocampal neurons.^2,132^ This dysregulation compromises mitochondrial bioenergetics and ATP production, exacerbating oxidative stress and triggering pro-apoptotic pathways, contributing to neuronal and motor dysfunction.^2^

Additionally, calcium homeostasis disruptions extend to the neuromuscular junction, where precise calcium signaling is essential for effective excitation-contraction coupling. APOE-*ε4*-associated calcium dysregulation may impair the timing and strength of muscle contractions, further exacerbating motor unit recruitment and synchronization deficits. In our study, blood calcium levels were marginally lower in APOE-*ε4* carriers compared to noncarriers (p = 0.05).

While blood calcium levels provide limited insight into intracellular calcium ion dynamics, this finding suggests a potential systemic effect of APOE-*ε4* on calcium homeostasis, warranting further investigation.

### 4.4 Neuronal hyperexcitability and neuromuscular compensation

Given the role of APOE-*ε4* in mitochondrial dysfunction, oxidative stress, and cellular metabolism, and their connection to muscle fatigue, the altered muscle activity observed in APOE-*ε4* carriers may represent a compensatory response to sustain function despite energy deficits and neuromuscular inefficiencies. Supporting this, previous research indicates that iPSC-derived neurons from APOE-*ε4* carriers exhibit higher excitatory current frequencies compared to isogenic APOE-*ε3* control neurons.^133,134^ This hyperexcitability may further disrupt motor function by altering the balance between excitatory and inhibitory signaling within motor circuits. Together, these metabolic and neuromuscular changes likely contribute to both increased muscle fatigue and adaptive shifts in motor unit behavior.

### 4.5 EMG-based neuromuscular markers: a potential risk indicator for AD

APOE-*ε4*-driven changes in EMG measures, including increased motor unit recruitment and synchronization, may serve as potential neuromuscular risk factors for AD. These measures outperformed standard cognitive tests in distinguishing APOE-*ε4* carriers from noncarriers (Table 4 and Figure 4). Furthermore, while MMSE showed the best cognitive test accuracy, it was more influenced by age, unlike EMG metrics—especially those from lip muscles—which were less age-dependent. These findings support EMG-based markers as sensitive, age-resilient indicators of subclinical motor decline, with potential to enhance early AD detection before cognitive symptoms emerge.

### 4.6 Sex-dependent impact of APOE-ε4 on neuromuscular function

A sex-specific pattern emerged in craniofacial muscle connectivity (Figure 1): female carriers showed a global decrease, while male carriers exhibited an increase, relative to noncarriers. This divergence may reflect an interaction between sex and APOE-*ε4* genotype.

In general, female APOE-*ε4* carriers face a substantially higher risk of developing AD compared to male carriers. ^135^ Women with the APOE-*ε4* allele experience more severe AD progression, with notable differences in neurological, biomarker, and pathological markers.^136–138^ Compared to male carriers, female APOE-*ε4* carriers demonstrate: 1) accelerated cognitive and functional decline, with a greater likelihood of MCI and AD progression^136,138^; 2) a fourfold increased risk of AD between ages 65–75 compared to noncarriers^104,136,138^; 3) higher cerebrospinal fluid total tau levels, indicative of greater neuronal degeneration^136,137^; 4) more pronounced Aβ deposition, tau accumulation, and reduced glucose metabolism in key memory-related brain regions^137^; 5) faster rates of brain structural atrophy compared to male carriers^139^; and 6) greater disruptions in brain connectivity and metabolic function, contributing to their heightened vulnerability to AD pathology.^103,137^

The interaction between sex and APOE-*ε4* is likely driven by a complex interplay of genetic, hormonal,^140^ vascular,^141^ and metabolic,^104^ factors. Notably, female APOE-*ε4* carriers show greater mitochondrial dysfunction,^104^ which may drive motor unit loss or remodeling, weakening muscle coupling and reducing functional connectivity. This impact of APOE-*ε4* in males appears less pronounced. Their motor unit function is likely better preserved, providing a sufficient pool of intact motor units to maintain effective muscle coordination. Their increased functional connectivity may represent a compensatory strategy—enhancing synchronous muscle activation to boost system “stiffness” and offset fatigue-related declines in performance.

Sex-specific differences in functional connectivity may limit its utility in distinguishing APOE-*ε4* carriers from noncarriers. Opposing trends in males and females can cancel each other out when analyzed together, reducing the discriminatory power of this measure. This likely explains why functional connectivity (X4) has a near-zero coefficient in the penalized logistic regression model (Table 4). These findings highlight the importance of stratifying analyses by sex to better capture genotype-specific neuromuscular risk factors.

### 4.7 Differential effects of APOE-ε4 on craniofacial musculature

This study reveals that APOE-*ε4* has a greater impact on facial rather than mandibular muscles. Facial muscles—especially those controlling lip movement—contain more slow-twitch fibers and exhibit greater degrees of freedom, while mandibular muscles are more robust, fast-twitch dominant.^142,143^ Additionally, facial muscles primarily receive contralateral cortical input, in contrast to the bilateral innervation of mandibular muscles.^144–146^ These anatomical and neurophysiological differences may make the facial musculature more vulnerable to APOE-*ε4*-related neuromuscular disruption, explaining the more pronounced effects observed.

It is also important to note that speech production requires moderate muscle activation, raising the question of whether the observed effects reflect APOE-*ε4* pathology or task-specific demands. In this study, the speech stimuli heavily featured bilabial plosives, requiring strong and coordinated activation of lip muscles, but also involving jaw opening and closing movements.

This may place a greater relative demand on lip muscles, potentially accelerating muscle fatigue and making them appear disproportionately affected by APOE-*ε4*. Consequently, while the observed pattern may partly reflect a task-induced effect, it remains unclear whether the greater involvement of facial versus mandibular muscles directly results from APOE-*ε4* pathology or is influenced by the motor demands of the task. Further studies utilizing varied speech and non-speech motor tasks are needed to disentangle APOE-*ε4*-related changes from task-driven effects.

### 4.8 Limitations and future directions

Future studies using a wider range of speech stimuli are needed to clarify the specific effects of APOE-*ε4* on orofacial muscles. In addition, Figure 3 shows considerable variability in EMG measures within each genotype group. While sex and agewere accounted for, other confounding factors may also contribute. Identifying these sources of variability will be key in future research.

## Consent Statement

Recruitment and consent procedures adhered to Mass General Brigham Healthcare (MGH) and HIPAA regulations. The study was approved by the MGB Institutional Review Board (IRB Protocol #2021P001460).

Before participating in testing, administrators thoroughly explained the experimental tasks and procedures, addressing any questions participants had. Relevant consent forms were provided, and each form was reviewed point by point with potential participants by a research staff member prior to obtaining their signature. Participants were explicitly informed of their right to withdraw from the study at any time during the experiment without any consequences and all participants provided written informed consent prior to participation.

## Data Availability

The raw speech recordings generated during the current study contain personally identifiable information and cannot be shared publicly for privacy and ethical reasons. A de-identified dataset comprising the extracted acoustic features that support the findings of this study is available from the corresponding author upon reasonable request.

## Author Contribution

M.E. served as the Principal Investigator and played a leading role in study conceptualization and design, data collection, data analysis, interpretation of findings, and manuscript writing. P.R. contributed to data analysis and manuscript writing. M.D. reviewed the manuscript and provided feedback on data analysis and statistical methodology. H.S. assisted with the annotation of audio signals aligned with EMG recordings. B.D.R. supported data collection, contributed to IRB submission and approval, reviewed the manuscript, and provided feedback. N.V.B. assisted with data collection, reviewed the manuscript, and provided feedback. D.H.S. contributed to participant characterization, manuscript review, and feedback. S.E.A. was involved in participant characterization and also reviewed and provided feedback on the manuscript. J.R.G. contributed to study conceptualization and design, interpretation of findings, and also reviewed the manuscript, and provided feedback. All authors contributed to the article and approved the submitted version.

## Bibliography

1. Mayeux R, Stern Y, Ottman R, et al. The apolipoprotein ε4 allele in patients with Alzheimer’s disease. Ann Neurol. 1993;34(5):752–754. doi:10.1002/ana.410340527

2. Pires M, Rego AC. Apoe4 and Alzheimer’s Disease Pathogenesis—Mitochondrial Deregulation and Targeted Therapeutic Strategies. Int J Mol Sci. 2023;24(1):778. doi:10.3390/ijms24010778

3. M. Di Battista A, M. Heinsinger N, William Rebeck G. Alzheimer’s Disease Genetic Risk Factor APOE-ε4 Also Affects Normal Brain Function. Curr Alzheimer Res. 2016;13(11):1200–1207. doi:10.2174/1567205013666160401115127

4. Tachibana M, Holm ML, Liu CC, et al. APOE4-mediated amyloid-β pathology depends on its neuronal receptor LRP1. J Clin Invest. 2019;129(3):1272–1277. doi:10.1172/JCI124853

5. Ferrari-Souza JP, Bellaver B, Ferreira PCL, et al. APOEε4 potentiates amyloid β effects on longitudinal tau pathology. Nat Aging. 2023;3(10):1210–1218. doi:10.1038/s43587-023-00490-2

6. Hampel H, Hardy J, Blennow K, et al. The Amyloid-β Pathway in Alzheimer’s Disease. Mol Psychiatry. 2021;26(10):5481–5503. doi:10.1038/s41380-021-01249-0

7. Brandon JA, Farmer BC, Williams HC, Johnson LA. APOE and Alzheimer’s Disease: Neuroimaging of Metabolic and Cerebrovascular Dysfunction. Front Aging Neurosci. 2018;10:180. doi:10.3389/fnagi.2018.00180

8. Therriault J, Benedet AL, Pascoal TA, et al. Association of Apolipoprotein E ε4 With Medial Temporal Tau Independent of Amyloid-β. JAMA Neurol. 2020;77(4):470. doi:10.1001/jamaneurol.2019.4421

9. Therriault J, Benedet AL, Pascoal TA, et al. APOEε4 potentiates the relationship between amyloid-β and tau pathologies. Mol Psychiatry. 2021;26(10):5977–5988. doi:10.1038/s41380-020-0688-6

10. Li W, Li R, Yan S, et al. Effect of *APOE ε* 4 genotype on amyloid-β, glucose metabolism, and gray matter volume in cognitively normal individuals and amnestic mild cognitive impairment. Eur J Neurol. 2023;30(3):587–596. doi:10.1111/ene.15656

11. Langbaum JBS, Chen K, Caselli RJ, et al. Hypometabolism in Alzheimer-Affected Brain Regions in Cognitively Healthy Latino Individuals Carrying the Apolipoprotein E ε4 Allele. Arch Neurol. 2010;67(4). doi:10.1001/archneurol.2010.30

12. Thambisetty M, Beason-Held L, An Y, Kraut MA, Resnick SM. APOE ε4 Genotype and Longitudinal Changes in Cerebral Blood Flow in Normal Aging. Arch Neurol. 2010;67(1). doi:10.1001/archneurol.2009.913

13. Rawle MJ, Davis D, Bendayan R, Wong A, Kuh D, Richards M. Apolipoprotein-E (Apoe) ε4 and cognitive decline over the adult life course. Transl Psychiatry. 2018;8(1):18. doi:10.1038/s41398-017-0064-8

14. Bretsky P, Guralnik JM, Launer L, Albert M, Seeman TE. The role of *APOE* -ε4 in longitudinal cognitive decline: MacArthur Studies of Successful Aging. Neurology. 2003;60(7):1077–1081. doi:10.1212/01.WNL.0000055875.26908.24

15. Mormino EC, Betensky RA, Hedden T, et al. Amyloid and *APOE ε4* interact to influence short-term decline in preclinical Alzheimer disease. Neurology. 2014;82(20):1760–1767. doi:10.1212/WNL.0000000000000431

16. Koutsodendris N, Blumenfeld J, Agrawal A, et al. Neuronal APOE4 removal protects against tau-mediated gliosis, neurodegeneration and myelin deficits. Nat Aging. 2023;3(3):275–296. doi:10.1038/s43587-023-00368-3

17. Zhang L, Xia Y, Gui Y. Neuronal ApoE4 in Alzheimer’s disease and potential therapeutic targets. Front Aging Neurosci. 2023;15:1199434. doi:10.3389/fnagi.2023.1199434

18. Parhizkar S, Holtzman DM. APOE mediated neuroinflammation and neurodegeneration in Alzheimer’s disease. Semin Immunol. 2022;59:101594. doi:10.1016/j.smim.2022.101594

19. Shi Y, Yamada K, Liddelow SA, et al. ApoE4 markedly exacerbates tau-mediated neurodegeneration in a mouse model of tauopathy. Nature. 2017;549(7673):523–527. doi:10.1038/nature24016

20. Friedberg JS, Aytan N, Cherry JD, et al. Associations between brain inflammatory profiles and human neuropathology are altered based on apolipoprotein E ε4 genotype. Sci Rep. 2020;10(1):2924. doi:10.1038/s41598-020-59869-5

21. Koutsodendris N, Blumenfeld J, Agrawal A, et al. APOE4-promoted gliosis and degeneration in tauopathy are ameliorated by pharmacological inhibition of HMGB1 release. Cell Rep. 2023;42(10):113252. doi:10.1016/j.celrep.2023.113252

22. Martins CAR, Oulhaj A, De Jager CA, Williams JH. *APOE* alleles predict the rate of cognitive decline in Alzheimer disease: A nonlinear model. Neurology. 2005;65(12):1888–1893. doi:10.1212/01.wnl.0000188871.74093.12

23. Morrison C, Oliver MD, Berry V, Kamal F, Dadar M. The influence of APOE status on rate of cognitive decline. GeroScience. 2024;46(3):3263–3274. doi:10.1007/s11357-024-01069-4

24. Whitehair DC, Sherzai A, Emond J, et al. Influence of apolipoprotein E ɛ4 on rates of cognitive and functional decline in mild cognitive impairment. Alzheimers Dement. 2010;6(5):412–419. doi:10.1016/j.jalz.2009.12.003

25. Albert M, Soldan A, Gottesman R, et al. Cognitive Changes Preceding Clinical Symptom Onset of Mild Cognitive Impairment and Relationship to ApoE Genotype. Curr Alzheimer Res. 2014;11(8):773–784. doi:10.2174/156720501108140910121920

26. Bunce D, Fratiglioni L, Small BJ, Winblad B, Bäckman L. APOE and cognitive decline in preclinical Alzheimer disease and non-demented aging. Neurology. 2004;63(5):816–821. doi:10.1212/01.WNL.0000137041.86153.42

27. Qian J, Betensky RA, Hyman BT, Serrano-Pozo A. Association of *APOE* Genotype With Heterogeneity of Cognitive Decline Rate in Alzheimer Disease. Neurology. 2021;96(19). doi:10.1212/WNL.0000000000011883

28. Caselli RJ, Dueck AC, Osborne D, et al. Longitudinal Modeling of Age-Related Memory Decline and the *APOE* ε4 Effect. N Engl J Med. 2009;361(3):255–263. doi:10.1056/NEJMoa0809437

29. Buchman AS, Boyle PA, Wilson RS, Beck TL, Kelly JF, Bennett DA. Apolipoprotein E e4 Allele is Associated With More Rapid Motor Decline in Older Persons. Alzheimer Dis Assoc Disord. 2009;23(1):63–69. doi:10.1097/WAD.0b013e31818877b5

30. Al-Chalabi A, Bakker MC, Shaw CE, et al. Association of apolipoprotein E &#x26;isin;4 allele with bulbar-onset motor neuron disease. The Lancet. 1996;347(8995):159–160. doi:10.1016/S0140-6736(96)90343-8

31. Sakurai R, Pieruccini-Faria F, Cornish B, et al. Link among *apolipoprotein E* E4, gait, and cognition in neurodegenerative diseases: ONDRI study. Alzheimers Dement. 2024;20(4):2968–2979. doi:10.1002/alz.13740

32. Jo S, Kim SO, Park KW, Lee SH, Hwang YS, Chung SJ. The role of APOE in cognitive trajectories and motor decline in Parkinson’s disease. Sci Rep. 2021;11(1):7819. doi:10.1038/s41598-021-86483-w

33. Garcia-Segura ME, Fischer CE, Schweizer TA, Munoz DG. APOE ɛ4/ɛ4 Is Associated with Aberrant Motor Behavior Through Both Lewy Body and Cerebral Amyloid Angiopathy Pathology in High Alzheimer’s Disease Pathological Load. J Alzheimers Dis. 2019;72(4):1077–1087. doi:10.3233/JAD-190643

34. Sakurai R, Montero-Odasso M. Apolipoprotein E4 Allele and Gait Performance in Mild Cognitive Impairment: Results From the Gait and Brain Study. J Gerontol Ser A. 2017;72(12):1676–1682. doi:10.1093/gerona/glx075

35. Choi HY, Liu Y, Tennert C, et al. APP interacts with LRP4 and agrin to coordinate the development of the neuromuscular junction in mice. eLife. 2013;2:e00220. doi:10.7554/eLife.00220

36. Maranzano A, Verde F, Dubini A, et al. Association of *APOE* genotype and cerebrospinal fluid Aβ and tau biomarkers with cognitive and motor phenotype in amyotrophic lateral sclerosis. Eur J Neurol. 2024;31(9):e16374. doi:10.1111/ene.16374

37. Yin J, Reiman EM, Beach TG, et al. Effect of ApoE isoforms on mitochondria in Alzheimer disease. Neurology. 2020;94(23). doi:10.1212/WNL.0000000000009582

38. Rong P, Heidrick L, Pattee G. A novel muscle network approach for objective assessment and profiling of bulbar involvement in ALS. Front Neurosci. 2025;18:1491997. doi:10.3389/fnins.2024.1491997

39. Rong P, Jawdat O. A novel physiologic marker of bulbar motor involvement in amyotrophic lateral sclerosis: Jaw muscle synergy. Clin Neurophysiol. 2021;132(1):94–103. doi:10.1016/j.clinph.2020.09.030

40. Rong P, Pattee GL. A potential upper motor neuron measure of bulbar involvement in amyotrophic lateral sclerosis using jaw muscle coherence. Amyotroph Lateral Scler Front Degener. 2021;22(5-6):368–379. doi:10.1080/21678421.2021.1874993

41. Yin J, Nielsen M, Carcione T, Li S, Shi J. Apolipoprotein E regulates mitochondrial function through the PGC-1α-sirtuin 3 pathway. Aging. 2019;11(23):11148–11156. doi:10.18632/aging.102516

42. Butterfield DA, Mattson MP. Apolipoprotein E and oxidative stress in brain with relevance to Alzheimer’s disease. Neurobiol Dis. 2020;138:104795. doi:10.1016/j.nbd.2020.104795

43. Fang W, Xiao N, Zeng G, et al. APOE4 genotype exacerbates the depression-like behavior of mice during aging through ATP decline. Transl Psychiatry. 2021;11(1):507. doi:10.1038/s41398-021-01631-0

44. Moore CA, Smith A, Ringel RL. Task-Specific Organization of Activity in Human Jaw Muscles. J Speech Lang Hear Res. 1988;31(4):670–680. doi:10.1044/jshr.3104.670

45. Nasreddine ZS, Phillips NA, Bédirian V, et al. The Montreal Cognitive Assessment, MoCA: A Brief Screening Tool For Mild Cognitive Impairment. J Am Geriatr Soc. 2005;53(4):695–699. doi:10.1111/j.1532-5415.2005.53221.x

46. Hoops S, Nazem S, Siderowf AD, et al. Validity of the MoCA and MMSE in the detection of MCI and dementia in Parkinson disease. Neurology. 2009;73(21):1738–1745. doi:10.1212/WNL.0b013e3181c34b47

47. Rossetti HC, Lacritz LH, Cullum CM, Weiner MF. Normative data for the Montreal Cognitive Assessment (MoCA) in a population-based sample. Neurology. 2011;77(13):1272–1275. doi:10.1212/WNL.0b013e318230208a

48. Freitas S, Simões MR, Alves L, Santana I. Montreal Cognitive Assessment: Validation Study for Mild Cognitive Impairment and Alzheimer Disease. Alzheimer Dis Assoc Disord. 2013;27(1):37–43. doi:10.1097/WAD.0b013e3182420bfe

49. Luis CA, Keegan AP, Mullan M. Cross validation of the Montreal Cognitive Assessment in community dwelling older adults residing in the Southeastern US. Int J Geriatr Psychiatry. 2009;24(2):197–201. doi:10.1002/gps.2101

50. Smith T, Gildeh N, Holmes C. The Montreal Cognitive Assessment: Validity and Utility in a Memory Clinic Setting. Can J Psychiatry. 2007;52(5):329–332. doi:10.1177/070674370705200508

51. Folstein MF, Folstein SE, McHugh PR. “Mini-mental state.” J Psychiatr Res. 1975;12(3):189–198. doi:10.1016/0022-3956(75)90026-6

52. Arevalo-Rodriguez I, Smailagic N, Roqué-Figuls M, et al. Mini-Mental State Examination (MMSE) for the early detection of dementia in people with mild cognitive impairment (MCI). Cochrane Dementia and Cognitive Improvement Group, ed. Cochrane Database Syst Rev. 2021;2021(7). doi:10.1002/14651858.CD010783.pub3

53. Tombaugh TN, McIntyre NJ. The Mini-Mental State Examination: A Comprehensive Review. J Am Geriatr Soc. 1992;40(9):922–935. doi:10.1111/j.1532-5415.1992.tb01992.x

54. Crum RM. Population-Based Norms for the Mini-Mental State Examination by Age and Educational Level. JAMA J Am Med Assoc. 1993;269(18):2386. doi:10.1001/jama.1993.03500180078038

55. Anthony JC, LeResche L, Niaz U, Von Korff MR, Folstein MF. Limits of the ‘Mini-Mental State’ as a screening test for dementia and delirium among hospital patients. Psychol Med. 1982;12(2):397–408. doi:10.1017/S0033291700046730

56. Holsinger T, Deveau J, Boustani M, Williams JW. Does This Patient Have Dementia? JAMA. 2007;297(21):2391. doi:10.1001/jama.297.21.2391

57. Bravo G, Hébert R. Age- and education-specific reference values for the Mini-Mental and Modified Mini-Mental State Examinations derived from a non-demented elderly population. Int J Geriatr Psychiatry. 1997;12(10):1008–1018. doi:10.1002/(SICI)1099-1166(199710)12:10<1008::AID-GPS676>3.0.CO;2-A

58. Bowie CR, Harvey PD. Administration and interpretation of the Trail Making Test. Nat Protoc. 2006;1(5):2277–2281. doi:10.1038/nprot.2006.390

59. Corrigan JD, Hinkeldey NS. Relationships between Parts A and B of the Trail Making Test. J Clin Psychol. 1987;43(4):402–409. doi:10.1002/1097-4679(198707)43:4<402::AID-JCLP2270430411>3.0.CO;2-E

60. Giovagnoli AR, Del Pesce M, Mascheroni S, Simoncelli M, Laiacona M, Capitani E. Trail making test: normative values from 287 normal adult controls. Ital J Neurol Sci. 1996;17(4):305–309. doi:10.1007/BF01997792

61. Linari I, Juantorena GE, Ibáñez A, Petroni A, Kamienkowski JE. Unveiling Trail Making Test: visual and manual trajectories indexing multiple executive processes. Sci Rep. 2022;12(1):14265. doi:10.1038/s41598-022-16431-9

62. Misdraji EL, Gass CS. The Trail Making Test and its neurobehavioral components. J Clin Exp Neuropsychol. 2010;32(2):159–163. doi:10.1080/13803390902881942

63. Muir RT, Lam B, Honjo K, et al. Trail Making Test Elucidates Neural Substrates of Specific Poststroke Executive Dysfunctions. Stroke. 2015;46(10):2755–2761. doi:10.1161/STROKEAHA.115.009936

64. Sánchez-Cubillo I, Periáñez JA, Adrover-Roig D, et al. Construct validity of the Trail Making Test: Role of task-switching, working memory, inhibition/interference control, and visuomotor abilities. J Int Neuropsychol Soc. 2009;15(3):438–450. doi:10.1017/S1355617709090626

65. Allen MD, Owens TE, Fong AK, Richards DR. A Functional Neuroimaging Analysis of the Trail Making Test-B: Implications for Clinical Application. Behav Neurol. 2011;24(2):159–171. doi:10.1155/2011/476893

66. Arbuthnott K, Frank J. Trail Making Test, Part B as a Measure of Executive Control: Validation Using a Set-Switching Paradigm. J Clin Exp Neuropsychol. 2000;22(4):518–528. doi:10.1076/1380-3395(200008)22:4;1-0;FT518

67. Gaudino EA, Geisler MW, Squires NK. Construct validity in the trail making test: What makes part B harder? J Clin Exp Neuropsychol. 1995;17(4):529–535. doi:10.1080/01688639508405143

68. Kortte KB, Horner MD, Windham WK. The Trail Making Test, Part B: Cognitive Flexibility or Ability to Maintain Set? Appl Neuropsychol. 2002;9(2):106–109. doi:10.1207/S15324826AN0902_5

69. MacPherson SE, Cox SR, Dickie DA, et al. Processing speed and the relationship between Trail Making Test-B performance, cortical thinning and white matter microstructure in older adults. Cortex. 2017;95:92–103. doi:10.1016/j.cortex.2017.07.021

70. Brandt J. The hopkins verbal learning test: Development of a new memory test with six equivalent forms. Clin Neuropsychol. 1991;5(2):125–142. doi:10.1080/13854049108403297

71. Benedict RHB, Schretlen D, Groninger L, Brandt J. Hopkins Verbal Learning Test – Revised: Normative Data and Analysis of Inter-Form and Test-Retest Reliability. Clin Neuropsychol. 1998;12(1):43–55. doi:10.1076/clin.12.1.43.1726

72. Shapiro AM, Benedict RHB, Schretlen D, Brandt J. Construct and Concurrent Validity of the Hopkins Verbal Learning Test – Revised. Clin Neuropsychol. 1999;13(3):348–358. doi:10.1076/clin.13.3.348.1749

73. Uttl B. Measurement of Individual Differences: Lessons From Memory Assessment in Research and Clinical Practice. Psychol Sci. 2005;16(6):460–467. doi:10.1111/j.0956-7976.2005.01557.x

74. Ross T, Calhoun E, Cox T, Wenner C, Kono W, Pleasant M. The reliability and validity of qualitative scores for the Controlled Oral Word Association Test. Arch Clin Neuropsychol. 2007;22(4):475–488. doi:10.1016/j.acn.2007.01.026

75. Benton AL. Differential behavioral effects in frontal lobe disease. Neuropsychologia. 1968;6(1):53–60. doi:10.1016/0028-3932(68)90038-9

76. Rodriguez-Aranda C, Martinussen M. Age-Related Differences in Performance of Phonemic Verbal Fluency Measured by Controlled Oral Word Association Task (COWAT): A Meta-Analytic Study. Dev Neuropsychol. 2006;30(2):697–717. doi:10.1207/s15326942dn3002_3

77. Ross TP. The reliability of cluster and switch scores for the controlled oral word association test. Arch Clin Neuropsychol. 2003;18(2):153–164. doi:10.1093/arclin/18.2.153

78. Bauer K, Malek-Ahmadi M. Meta-analysis of Controlled Oral Word Association Test (COWAT) FAS performance in amnestic mild cognitive impairment and cognitively unimpaired older adults. Appl Neuropsychol Adult. 2023;30(4):424–430. doi:10.1080/23279095.2021.1952590

79. Henry JD, Crawford JR. Verbal fluency deficits in Parkinson’s disease: A meta-analysis. J Int Neuropsychol Soc. 2004;10(4):608–622. doi:10.1017/S1355617704104141

80. Gladsjo JA, Schuman CC, Evans JD, Peavy GM, Miller SW, Heaton RK. Norms for Letter and Category Fluency: Demographic Corrections for Age, Education, and Ethnicity. Assessment. 1999;6(2):147–178. doi:10.1177/107319119900600204

81. Benton AL, Hamsher K deS, Sivan AB. Multilingual Aphasia Examination. AJA associates; 1994.

82. Lezak MD, Hannay HJ, Howieson DB, Loring DW. Neuropsychological Assessment. 4. ed. Oxford Univ. Press; 2004.

83. Steinberg BA, Bieliauskas LA, Smith GE, Ivnik RJ. Mayo’s Older Americans Normative Studies: Age-and IQ-Adjusted Norms for the Trail-Making Test, the Stroop Test, and MAE Controlled Oral Word Association Test. Clin Neuropsychol. 2005;19(3-4):329–377. doi:10.1080/13854040590945210

84. Heaton RK, Grant I, Matthews CG. Comprehensive Norms for an Expanded Halstead-Reitan Battery: Demographic Corrections, Research Findings, and Clinical Applications. Psychological Assessment Resources; 1991. https://books.google.com/books?id=S47HHJLrMtIC

85. Troyer AK, Moscovitch M, Winocur G. Clustering and switching as two components of verbal fluency: Evidence from younger and older healthy adults. Neuropsychology. 1997;11(1):138–146. doi:10.1037/0894-4105.11.1.138

86. Strauss EH, Spreen O, Sherman EMS. A Compendium of Neuropsychological Tests: Administration, Norms, and Commentary. Third edition. Oxford University Press; 2006.

87. The MathWorks Inc. MATLAB version: 9.14.0 (R2023a). Published online 2023. https://www.mathworks.com

88. De Luca CJ, Donald Gilmore L, Kuznetsov M, Roy SH. Filtering the surface EMG signal: Movement artifact and baseline noise contamination. J Biomech. 2010;43(8):1573–1579. doi:10.1016/j.jbiomech.2010.01.027

89. Stepp CE. Surface Electromyography for Speech and Swallowing Systems: Measurement, Analysis, and Interpretation. J Speech Lang Hear Res. 2012;55(4):1232–1246. doi:10.1044/1092-4388(2011/11-0214)

90. Rong P, Pattee GL. A multidimensional facial surface EMG analysis for objective assessment of bulbar involvement in amyotrophic lateral sclerosis. Clin Neurophysiol. 2022;135:74–84. doi:10.1016/j.clinph.2021.11.074

91. Yaar I, Niles L. Muscle fiber conduction velocity and mean power spectrum frequency in neuromuscular disorders and in fatigue. Muscle Nerve. 1992;15(7):780–787. doi:10.1002/mus.880150706

92. Halliday DM, Rosenberg JR, Amjad AM, Breeze P, Conway BA, Farmer SF. A framework for the analysis of mixed time series/point process data—Theory and application to the study of physiological tremor, single motor unit discharges and electromyograms. Prog Biophys Mol Biol. 1995;64(2-3):237–278. doi:10.1016/S0079-6107(96)00009-0

93. Terry K, Griffin L. How computational technique and spike train properties affect coherence detection. J Neurosci Methods. 2008;168(1):212–223. doi:10.1016/j.jneumeth.2007.09.014

94. Boonstra TW, Danna-Dos-Santos A, Xie HB, Roerdink M, Stins JF, Breakspear M. Muscle networks: Connectivity analysis of EMG activity during postural control. Sci Rep. 2015;5(1):17830. doi:10.1038/srep17830

95. Brown P. Cortical drives to human muscle: the Piper and related rhythms. Prog Neurobiol. 2000;60(1):97–108. doi:10.1016/S0301-0082(99)00029-5

96. Laine CM, Martinez-Valdes E, Falla D, Mayer F, Farina D. Motor Neuron Pools of Synergistic Thigh Muscles Share Most of Their Synaptic Input. J Neurosci. 2015;35(35):12207–12216. doi:10.1523/JNEUROSCI.0240-15.2015

97. Hudgins B, Parker P, Scott RN. A new strategy for multifunction myoelectric control. IEEE Trans Biomed Eng. 1993;40(1):82–94. doi:10.1109/10.204774

98. Rioul O, Vetterli M. Wavelets and signal processing. IEEE Signal Process Mag. 1991;8(4):14–38. doi:10.1109/79.91217

99. Eckmann JP, Kamphorst SO, Ruelle D. Recurrence Plots of Dynamical Systems. Europhys Lett EPL. 1987;4(9):973–977. doi:10.1209/0295-5075/4/9/004

100. Marwan N, Carmenromano M, Thiel M, Kurths J. Recurrence plots for the analysis of complex systems. Phys Rep. 2007;438(5-6):237–329. doi:10.1016/j.physrep.2006.11.001

101. R Core Team. R: A Language and Environment for Statistical Computing. R Foundation for Statistical Computing; 2021. https://www.R-project.org/

102. Kuhn M. Building Predictive Models in *R* Using the **caret** Package. J Stat Softw. 2008;28(5). doi:10.18637/jss.v028.i05

103. Sundermann EE, Tran M, Maki PM, Bondi MW, Alzheimer’s Disease Neuroimaging Initiative. Sex differences in the association between apolipoprotein E ε4 allele and Alzheimer’s disease markers. Alzheimers Dement Diagn Assess Dis Monit. 2018;10(1):438–447. doi:10.1016/j.dadm.2018.06.004

104. Arnold M, Nho K, Kueider-Paisley A, et al. Sex and APOE ε4 genotype modify the Alzheimer’s disease serum metabolome. Nat Commun. 2020;11(1):1148. doi:10.1038/s41467-020-14959-w

105. Damoiseaux JS, Seeley WW, Zhou J, et al. Gender Modulates the APOE 4 Effect in Healthy Older Adults: Convergent Evidence from Functional Brain Connectivity and Spinal Fluid Tau Levels. J Neurosci. 2012;32(24):8254–8262. doi:10.1523/JNEUROSCI.0305-12.2012

106. Yan S, Zheng C, Paranjpe MD, et al. Sex modifies APOE ε4 dose effect on brain tau deposition in cognitively impaired individuals. Brain. 2021;144(10):3201–3211. doi:10.1093/brain/awab160

107. Gamache J, Yun Y, Chiba-Falek O. Sex-dependent effect of *APOE* on Alzheimer’s disease and other age-related neurodegenerative disorders. Dis Model Mech. 2020;13(8):dmm045211. doi:10.1242/dmm.045211

108. Revelle W. Psych: Procedures for Psychological, Psychometric, and Personality Research. Northwestern University; 2024. https://CRAN.R-project.org/package=psych

109. Berisha V, Krantsevich C, Hahn PR, et al. Digital medicine and the curse of dimensionality. Npj Digit Med. 2021;4(1):153. doi:10.1038/s41746-021-00521-5

110. Panzarella E, Beribisky N, Cribbie RA. Denouncing the use of field-specific effect size distributions to inform magnitude. PeerJ. 2021;9:e11383. doi:10.7717/peerj.11383

111. Farina D, Fattorini L, Felici F, Filligoi G. Nonlinear surface EMG analysis to detect changes of motor unit conduction velocity and synchronization. J Appl Physiol. 2002;93(5):1753–1763. doi:10.1152/japplphysiol.00314.2002

112. Hagg GM. Interpretation of EMG spectral alterations and alteration indexes at sustained contraction. J Appl Physiol. 1992;73(4):1211–1217. doi:10.1152/jappl.1992.73.4.1211

113. Wan J jing, Qin Z, Wang P yuan, Sun Y, Liu X. Muscle fatigue: general understanding and treatment. Exp Mol Med. 2017;49(10):e384–e384. doi:10.1038/emm.2017.194

114. Husain MA, Laurent B, Plourde M. APOE and Alzheimer’s Disease: From Lipid Transport to Physiopathology and Therapeutics. Front Neurosci. 2021;15:630502. doi:10.3389/fnins.2021.630502

115. Yin F. Lipid metabolism and Alzheimer’s disease: clinical evidence, mechanistic link and therapeutic promise. FEBS J. 2023;290(6):1420–1453. doi:10.1111/febs.16344

116. Contessa P, De Luca CJ, Kline JC. The compensatory interaction between motor unit firing behavior and muscle force during fatigue. J Neurophysiol. 2016;116(4):1579–1585. doi:10.1152/jn.00347.2016

117. Amann M, Sidhu SK, Weavil JC, Mangum TS, Venturelli M. Autonomic responses to exercise: Group III/IV muscle afferents and fatigue. Auton Neurosci. 2015;188:19–23. doi:10.1016/j.autneu.2014.10.018

118. Pethick J, Tallent J. The Neuromuscular Fatigue-Induced Loss of Muscle Force Control. Sports. 2022;10(11):184. doi:10.3390/sports10110184

119. Lyon P, Strippoli V, Fang B, Cimmino L. B Vitamins and One-Carbon Metabolism: Implications in Human Health and Disease. Nutrients. 2020;12(9):2867. doi:10.3390/nu12092867

120. Ashok T, Puttam H, Tarnate VCA, et al. Role of Vitamin B12 and Folate in Metabolic Syndrome. Cureus. Published online October 6, 2021. doi:10.7759/cureus.18521

121. Finsterer J. Biomarkers of peripheral muscle fatigue during exercise. BMC Musculoskelet Disord. 2012;13(1):218. doi:10.1186/1471-2474-13-218

122. Quaye IK. Extracellular hemoglobin: the case of a friend turned foe. Front Physiol. 2015;6. doi:10.3389/fphys.2015.00096

123. Johnson CN, Lysaker CR, McCoin CS, et al. Skeletal muscle proteome differs between young APOE3 and APOE4 targeted replacement mice in a sex-dependent manner. Front Aging Neurosci. 2024;16:1486762. doi:10.3389/fnagi.2024.1486762

124. Zhang J, Zhang Y, Wang J, Xia Y, Zhang J, Chen L. Recent advances in Alzheimer’s disease: mechanisms, clinical trials and new drug development strategies. Signal Transduct Target Ther. 2024;9(1):211. doi:10.1038/s41392-024-01911-3

125. Huang J, Li C, Shang H. Astrocytes in Neurodegeneration: Inspiration From Genetics. Front Neurosci. 2022;16:882316. doi:10.3389/fnins.2022.882316

126. Houldsworth A. Role of oxidative stress in neurodegenerative disorders: a review of reactive oxygen species and prevention by antioxidants. Brain Commun. 2023;6(1):pfcad356. doi:10.1093/braincomms/fcad356

127. Ramassamy C, Averill D, Beffert U, et al. Oxidative Insults Are Associated with Apolipoprotein E Genotype in Alzheimer’s Disease Brain. Neurobiol Dis. 2000;7(1):23–37. doi:10.1006/nbdi.1999.0273

128. D’Amico E, Factor-Litvak P, Santella RM, Mitsumoto H. Clinical perspective on oxidative stress in sporadic amyotrophic lateral sclerosis. Free Radic Biol Med. 2013;65:509–527. doi:10.1016/j.freeradbiomed.2013.06.029

129. Fernandez CG, Hamby ME, McReynolds ML, Ray WJ. The Role of APOE4 in Disrupting the Homeostatic Functions of Astrocytes and Microglia in Aging and Alzheimer’s Disease. Front Aging Neurosci. 2019;11:14. doi:10.3389/fnagi.2019.00014

130. Foley KE, Diemler CA, Hewes AA, Garceau DT, Sasner M, Howell GR. *APOE* ε4 and exercise interact in a sex-specific manner to modulate dementia risk factors. Alzheimers Dement Transl Res Clin Interv. 2022;8(1):e12308. doi:10.1002/trc2.12308

131. Chaudhari K, Wong JM, Vann PH, Como T, O’Bryant SE, Sumien N. ApoE Genotype-Dependent Response to Antioxidant and Exercise Interventions on Brain Function. Antioxidants. 2020;9(6):553. doi:10.3390/antiox9060553

132. Liang T, Hang W, Chen J, et al. ApoE4 (Δ272–299) induces mitochondrial-associated membrane formation and mitochondrial impairment by enhancing GRP75-modulated mitochondrial calcium overload in neuron. Cell Biosci. 2021;11(1):50. doi:10.1186/s13578-021-00563-y

133. Nuriel T, Angulo SL, Khan U, et al. Neuronal hyperactivity due to loss of inhibitory tone in APOE4 mice lacking Alzheimer’s disease-like pathology. Nat Commun. 2017;8(1):1464. doi:10.1038/s41467-017-01444-0

134. Targa Dias Anastacio H, Matosin N, Ooi L. Neuronal hyperexcitability in Alzheimer’s disease: what are the drivers behind this aberrant phenotype? Transl Psychiatry. 2022;12(1):257. doi:10.1038/s41398-022-02024-7

135. Neu SC, Pa J, Kukull W, et al. Apolipoprotein E Genotype and Sex Risk Factors for Alzheimer Disease: A Meta-analysis. JAMA Neurol. 2017;74(10):1178. doi:10.1001/jamaneurol.2017.2188

136. O’Neal MA. Women and the risk of Alzheimer’s disease. Front Glob Womens Health. 2024;4:1324522. doi:10.3389/fgwh.2023.1324522

137. Mares J, Kumar G, Sharma A, et al. *APOE* ε4–associated heterogeneity of neuroimaging biomarkers across the Alzheimer’s disease continuum. Alzheimers Dement. 2025;21(1):e14392. doi:10.1002/alz.14392

138. Ungar L, Altmann A, Greicius MD. Apolipoprotein E, gender, and Alzheimer’s disease: an overlooked, but potent and promising interaction. Brain Imaging Behav. 2014;8(2):262–273. doi:10.1007/s11682-013-9272-x

139. Sampedro F, Vilaplana E, De Leon MJ, et al. *APOE* -by-sex interactions on brain structure and metabolism in healthy elderly controls. Oncotarget. 2015;6(29):26663–26674. doi:10.18632/oncotarget.5185

140. Raber J, Bongers G, LeFevour A, Buttini M, Mucke L. Androgens Protect against Apolipoprotein E4-Induced Cognitive Deficits. J Neurosci. 2002;22(12):5204–5209. doi:10.1523/JNEUROSCI.22-12-05204.2002

141. Shinohara M, Murray ME, Frank RD, et al. Impact of sex and APOE4 on cerebral amyloid angiopathy in Alzheimer’s disease. Acta Neuropathol (Berl). 2016;132(2):225–234. doi:10.1007/s00401-016-1580-y

142. Rowlerson A, Raoul G, Daniel Y, et al. Fiber-type differences in masseter muscle associated with different facial morphologies. Am J Orthod Dentofacial Orthop. 2005;127(1):37–46. doi:10.1016/j.ajodo.2004.03.025

143. Pradebon A, Pradebon M, Goulart G, et al. Biomechanics Potential of the Masseter and Temporal Muscles in the Mandibles of Mesofacial Subjects. J Morphol Sci. 2018;35(03):180–182. doi:10.1055/s-0038-1675569

144. Hylander W. Functional anatomy and biomechanics of the masticatory apparatus. Temporomandibular Disord Evidenced Approach Diagn Treat. Published online January 2006:1–34.

145. Committee on Temporomandibular Disorders (TMDs): From Research Discoveries to Clinical Treatment, Board on Health Sciences Policy, Board on Health Care Services, Health and Medicine Division, National Academies of Sciences, Engineering, and Medicine. Temporomandibular Disorders: Priorities for Research and Care. (Bond EC, Mackey S, English R, Liverman CT, Yost O, eds.). National Academies Press; 2020:25652. doi:10.17226/25652

146. Asadi H, Budenz A. Anatomy of the Masticatory System. In: Gremillion HA, Klasser GD, eds. Temporomandibular Disorders. Springer International Publishing; 2018:17–33. doi:10.1007/978-3-319-57247-5_2

